# Phase I Clinical Study of DOC1021 (dubodencel) for Adjuvant Immunotherapy of Glioblastoma

**DOI:** 10.64898/2026.03.28.26349013

**Authors:** JF Georges, C Clay, S Amin, A Goralczyk, C Mossop, CJ Bilbao, A Valeri, J Ifrach, M Zaher, L Colman, L Kohler, EH Schumann, M Vu, BA Burns, A Trivedi, W Liu, M Namekar, CJ Hofferek, KJ Ernste, N Bisht, J Vazquez-Perez, D Oyewole-Said, SB Amanya, V Rodriguez, DC Kraushaar, D Okoebor, I Bellayr, J Hartenbach, MM Halpert, EM Duus, LK Aguilar, SH Hsu, J-J Zhu, RC Zvavanjanja, Y Bai, SW Kang, H-J Jang, HS Lee, R Garg, Y Esquenazi, N Tandon, A Turtz, V Konduri, WK Decker

**Author notes:** These two authors contributed equally to the preparation of this manuscript (V Konduri, WK Decker). Earlier results of this study were presented at an oral session at American Society of Clinical Oncology annual meeting on May 31, 2025. *J Clin Oncol* **43**, 2014-2014(2025).

## Abstract

**PURPOSE:** Newly-diagnosed glioblastoma (nGBM) is a devastating tumor with median survival of only 14-18 months despite aggressive standard of care (SOC). Dendritic cell (DC) homologous antigenic double-loading provides a powerful pattern-based signal that initiates cDC1-like skewing of monocytic precursors, inducing downstream development of CD8^+^ memory effectors. Here we report phase I results for DOC1021 (dubodencel), a novel DC vaccine regimen integrated with SOC.

**METHODS:** In this dose-escalating study, DC prepared from mobilized peripheral blood were doubly loaded with autologous tumor lysate and amplified tumor mRNA and administered bilaterally near the deep cervical node chains in three biweekly courses given with weekly peg-IFN after conclusion of chemoradiation. Four dose levels from 3.5x10^6^ to 3.6x10^7^ total cells were tested. Patients with subtotal resection or tumor progression prior to vaccination were not excluded.

**RESULTS:** Eighteen patients (median age 61 years (range 47-73), 94% MGMT unmethylated, 25% subtotal/partial resected) completed vaccination (16 nGBM, 2 recurrent) with no dose-limiting toxicities. Attributable AE were mostly mild and flu-like or injection-site reactions. Twelve-month OS among the newly-diagnosed cohort was 88% compared to an expected ∼60% for SOC alone. Patients who received observation rather than reoperation in response to worsening MRI contrast-enhancement demonstrated gradual lesional resolution and improved OS. Immunophenotyping revealed post-vaccination elevations in CD4 and CD8 memory T-cells in peripheral blood, and spatial transcriptomic analysis revealed foci of activated inflammatory complexes at the primary tumor site.

**CONCLUSIONS:** DOC1021 was safe, feasibly integrated within SOC, and associated with more favorable outcomes in this challenging patient population. Patients who received observation rather than reoperation for worsening MRI contrast-enhancement exhibited superior survival, suggesting an immune-reactive tumor microenvironment manifesting as pseudo-progression. These data supported initiation of a randomized Phase II trial (NCT06805305) for nGBM.

## Introduction

Glioblastoma (GBM) is an aggressive, isocitrate dehydrogenase (IDH)-wildtype primary neoplasm of the central nervous system (CNS)[1]. Since the introduction of temozolomide chemotherapy in 2005 based on a 2.5 month improvement in median OS over radiotherapy alone[2], there have been no new drug approvals for the treatment of nGBM. Therefore, despite improvements in surgical techniques and local therapies such as tumor-treating fields (TTFs), GBM remains a devastating disease with median OS of 14-18 months and five-year OS of only 5-7%.[3, 4] While the extent of tumor resection is a critical prognostic factor[5], complete resection is not achievable due to the infiltrative nature of glioblastoma and the need to preserve eloquent brain regions. Additionally, the methylation status of the O^6^-methylguanine–DNA methyltransferase (MGMT) gene promoter is pivotal to prognosis and treatment response. MGMT plays a key role in DNA repair, and heightened repair capacity in MGMT unmethylated tumors diminishes the effectiveness of alkylating chemotherapy, leading to poorer outcomes.[6] For the 50-60% of GBM patients harboring an unmethylated MGMT promoter, median OS is only 12-14 months compared to 22-29 months among patients with MGMT methylated tumors.[7, 8] The dire clinical course of GBM, specifically MGMT unmethylated GBM, necessitates the development of novel therapeutic modalities.

Over the last two decades, immune potentiating interventions[9–13] have become standard approaches for the treatment of cancer. A broad spectrum of immunotherapies have also been evaluated in clinical trials for GBM including classical vaccines, dendritic cell therapies, chimeric antigen receptor (CAR) T-cells, checkpoint inhibitors, and oncolytic viruses.[8, 14–17] While CAR-T and oncolytic virus approaches remain works in progress, the majority of other regimens in this space have been unsuccessful, underscored by two recent Phase III checkpoint inhibitor failures likely due in part to the presence of multiple immunoinhibitory pathways that prevent anti-tumor T-cell activation.[18, 19] Moreover, glioblastomas are heterogeneous tumors, limiting the potential for approaches that target limited numbers of surface antigens and for which epitope loss can lead to resistance (i.e. as in EGFRvIII targeted approaches).[14, 16] Effective immunotherapeutic approaches to GBM will necessarily need to 1) target a multitude of tumor antigens, 2) easily permeate the blood-brain barrier, and 3) alter the immunosuppressive tumor microenvironment, a challenging combination of requirements Homologous antigenic loading of dendritic cells (DC) is a novel technology that leverages recent innovations in immunobiology. The loading of DC with substantially identical or “homologous” MHC class I and II peptide epitopes is a signal recognized as a critical viral pathogen-associated molecular pattern (PAMP), the detection of which leads to release of the AIMp1 signaling intermediate from a sensor complex known as the mARS.[20–22] AIMp1 release in turn initiates differentiation cascades through modification of the AP-1 heterodimeric transcription factor, catalyzing a reorganization of the DC transcriptome that produces an antigen-presenting cell type with cDC1-like properties.[23, 24] Stimulation of monocyte-derived DC in this manner also abrogates constitutive secretion of DC CTLA-4^+^ microvesicles[20, 25], favoring downstream development of high potency CD161^int^ CD8^+^ T-cells that exhibit an enhanced serial killing, resistance to exhaustion, and a tissue homing capacity ideal for the treatment of solid tumors[26–28] as demonstrated in preclinical model systems.[29, 30]

Here, we report results of a first-in-human phase I clinical trial of DOC1021 (dubodencel), an autologous DC vaccine generated through homologous antigenic loading using both patient-derived tumor lysate and *in vitro*-amplified tumor mRNA and delivered to interferon-adjuvanted patients by ultrasound-guided injection in the vicinity of the deep cervical node chains.

## Materials and Methods

### Study Design

Patients with GBM were enrolled in this single-arm phase I clinical trial (NCT04552886) between October 2021 and December 2023. This study was conducted at two sites, Cooper University Hospital and the University of Texas Health Science Center, Houston. The trial was approved by all necessary institutional review boards including the WCG institutional review board (reference #20202650) and conducted under an FDA Investigational New Drug application. All patients were provided with written consent prior to enrollment. Eligibility criteria are listed in the clinical protocol. Key inclusion criteria included adult patients diagnosed with IDH-wt GBM deemed to be potentially resectable and eligible for postoperative adjuvant chemoradiation; adequate kidney, liver, bone marrow, and immune function; and ECOG performance ≤ 2. Key exclusion criteria included any severe or uncontrolled medical condition that could affect study participation; use of non-standard post-operative treatment regimen; and concurrent/expected need for corticosteroids during the vaccination phase of the study. MGMT unmethylated was defined as < 15% MGMT promoter CpG island methylation as ascertained by local assay. Use of tumor-treating fields (TTF) and/or bevacizumab with adjuvant TMZ were permitted at the discretion of the treating investigator. For post-vaccination clinical follow up, patients were evaluated weekly by a neurooncologist during the 6-week period of vaccination and pegylated interferon alpha 2a (peg-IFN) administration. Weekly physical exams, complete metabolic panels and complete blood count labs were obtained to monitor for toxicity. Peripheral blood draws taken at baseline and at post-vaccination study day 106-108 were utilized to analyze patient peripheral immune responses by flow cytometry. Standard Bayesian optimal interval (BOIN) trial design was applied.[31]

### Patient Sample Processing

Perioperatively and following sufficient tissue acquisition for clinical assessments, a tissue sample of at least 200 mg was obtained from the patient. This sample was flash frozen in liquid nitrogen-based, GMP compliant dry shipper (Cryoport Systems, Irvine, CA) at -195° C. The sample was transported overnight to the laboratory for isolation of protein lysate and total RNA. Within four weeks of surgical resection, patients received daily recombinant granulocyte colony-stimulating factor (filgrastim G-CSF) injections for five days. Patients underwent leukapheresis on the fifth day of G-CSF administration to collect 1 x 10^10^ total nucleated cells (TNC). Apheresis products were then shipped to the manufacturing facility in a GMP-compliant room temperature shipper (Cryoport). Cell-based vaccines were prepared with autologous patient tumor lysate and amplified mRNA as described.[26]

### Vaccine Administration

Vaccination began within two weeks of completing SOC chemoradiation for newly-diagnosed GBM. Recurrent GBM patients could follow the same treatment protocol/timings as those newly diagnosed with GBM or alternatively undergo an abbreviated treatment schedule to initiate a more rapid implementation of the vaccination schedule at the discretion of the neuro-oncology team. On the day of vaccination, product was retrieved from liquid nitrogen storage and thawed in a 37° C bedside waterbath. Vaccine cells were diluted to a volume of 2.0 ml in medical grade saline and administered bilaterally by ultrasound-guided injection within 0.5 cm of the deep cervical lymph node chain. Patients received 180 mcg peg-IFN (Pegasys, Genentech, South San Francisco, CA) subcutaneously on the day of vaccination and continued to receive weekly peg-IFN injections for a total of six administrations. Dendritic cell vaccinations were repeated every other week for a total of three administrations. Dose escalation was utilized to interrogate the safety of 4 dose levels including 3.5 x 10^6^, 7 x 10^6^, 1.4 x 10^7^ and 3.6 x 10^7^ dendritic cells). Dose modifications for peg-IFN were specified in the protocol. A data safety monitoring board (DSMB) meeting occurred after at least three patients completed each dose level and prior to escalation to the next dose cohort.

### Magnetic Resonance Imaging Data Acquisition and Analysis

Magnetic resonance im-ages were obtained preoperatively and within 72 hours after tumor resection. Follow-up MRIs were performed approximately every 8 weeks as per standard of care or more frequently if progression was suspected. Extent of resection was determined by comparing the pre-operative to the post-operative (within 72 hours) MRI based on total contrast enhancing tumor volume and calculated as follows: (pre-operative tumor volume - post-operative tumor volume)/pre-operative tumor volume x 100%. Extent of resection categories were defined as follows: Gross total resection (GTR) 100% of contrast-enhancing (CE) tumor removed; Near total resection (NTR) ≥ 95% CE tumor removed and ≤ 1 cm^3^ residual; Subtotal resection (STR) ≥ 80% CE tumor removed and ≤ 5 cm^3^ residual; Partial resection < 80% or > 5 cm^3^ residual.

### Spatial Transcriptome Library Construction, Sequencing, and Cluster Generation

Tissue sections were mounted onto poly-L-lysine coated slides, deparaffinized, H&E stained, and imaged. One library per section was generated using the Visium CytAssist Spatial Gene Expression for FFPE kit according to the manufacturer’s instructions (10X Genomics, Pleasanton, CA). Briefly, sections were destained and decrosslinked prior to probe hybridization overnight. Following hybridization, probe pairs were ligated and tissues restained with H&E and imaged on the CytAssist instrument. Sections were treated with RNase prior to tissue permeabilization. Slides were loaded onto the CytAssist where probe release and transfer to the capture area (6.5 x 6.5 mm) of the 10X Visium GEX slide were performed. Capture probes were extended, pre-amplified, and the resulting PCR product purified. Probe libraries were indexed in a final PCR reaction with 12-15 cycles, as determined by the qPCR, prior to cleanup. The resulting libraries were pooled in equimolar fashion and each library was sequenced on a NovaSeq 6000 Sequencing System (Illumina) to an average depth of 116 million reads.

### Analysis of spatial transcriptomics data

Sequencing reads were aligned to the human transcriptome (GRCh38) using the Spaceranger count pipeline (10x Genomics, Spaceranger v3.0.0)[32]. Downstream analysis was conducted with SCANPY (v1.10.3)[33], supplemented by Python-based libraries including pandas, numpy, and seaborn for data manipulation and visualization. Low-quality cells were excluded based on stringent filtering criteria: cells with fewer than 200 detected genes or mitochondrial gene content below 1% were removed. This filtering ensured the retention of high-quality cells for further analysis. To leverage spatial information, tissue regions from GBM samples were separated into independent datasets. This segregation enabled precise and independent analysis of the spatial transcriptomic landscapes of each sample. For integrated analysis, datasets from multiple samples were merged into a unified dataset. Batch effects were addressed using the scVI method[34], which effectively mitigates technical variation across samples while preserving essential biological variability. This robust approach minimized inter-batch variability, ensuring reliable downstream analyses. The resulting segmentation data were integrated with alignment information to achieve comprehensive spatial mapping of cells. The Loupe Browser was utilized to modify specific layers of dot information, which were subsequently re-applied to the original data. This workflow provided a robust and detailed framework for analyzing spatial transcriptomics data, enabling insights into the complex spatial organization of the human GBM transcriptome.

### Immune Triad Analysis Using VISIUM^TM^ Spatial Transcriptomics

Spatial spots were deconvoluted using signature gene sets to identify spots enriched for activated CD4⁺ T cells (CD4⁺CD25⁺FOXP3⁻), activated CD8⁺ T cells (CD8a⁺CD25⁺), hematopoietic-derived microglial cells (CD68⁺C1QA⁺OLFML3⁺), and regulatory T cells (CD4⁺CD25⁺Foxp3⁺). Spots expressing markers of all three immune cell types (activated CD4⁺, activated CD8⁺, and microglia) were defined as “triad” spots. Treg-enriched and other immune-enriched spots were also annotated based on canonical marker expression. For each triad-enriched spot, a circular neighborhood with a 300 µm diameter was defined. Within each neighborhood, the density of surrounding triad and Treg spots was then calculated. Mean densities across samples were compared between pre- and post-vaccination using paired Wilcoxon signed-rank tests.

### Flow Cytometry

Cryopreserved PBMC were processed and stained using standard methods detailed in Supplemental methods. Data analysis was performed using FlowJo software v10.10 (BD).

### Statistical Analysis

Significance of differences, unless stated otherwise, was determined by one-way analysis of variance (ANOVA) using Tukey’s honestly significant difference (HSD) post hoc test. Pairwise comparisons were performed by Student’s two-tailed t-test. Kaplan–Meier survival significance was determined by the log rank (Mantel-Cox) test. All data are displayed as the mean ± SEM or SD as stated. Survival was estimated by Kaplan-Meier analysis. GBM-003 was censored at time of death from suicide, and 2 living patients were censored at date of last follow-up. All analyses were performed using Prism software version 10.3.1 for iOS (GraphPad Software, Boston, MA) or Microsoft Excel for Mac version 16.30. Statistical significance was defined as *p* ≤ 0.05. Data cutoff for survival analyses was November 1, 2025.

### Data Availability

The Visium spatial transcriptomic data that support the findings of this study are available in the Gene Expression Omnibus (GEO) at http://ncbi.nlm.nih.gov/geo, accession number GSE286477.

## Results

### Study Design and Patient Characteristics

A study schema is shown in **Figure 1A**. Between October 2021 and December 2023, 18 patients (16 newly diagnosed and 2 recurrent) completed the study treatment that included three bilateral administrations of DOC1021 with peg-IFN plus standard of care Stupp protocol. An additional patient discontinued after one DOC1021 dose due to non-compliance/patient choice. Nine patients consented but were not treated due to postsurgical decline in performance status (n=3), non-compliance/patient choice (n=2), other medical event (n=1), brainstem lesion (n=1), diagnosis not GBM (n=1) and manufacturing failure (n=1). Four dose levels were evaluated as planned.

**Figure 1.**
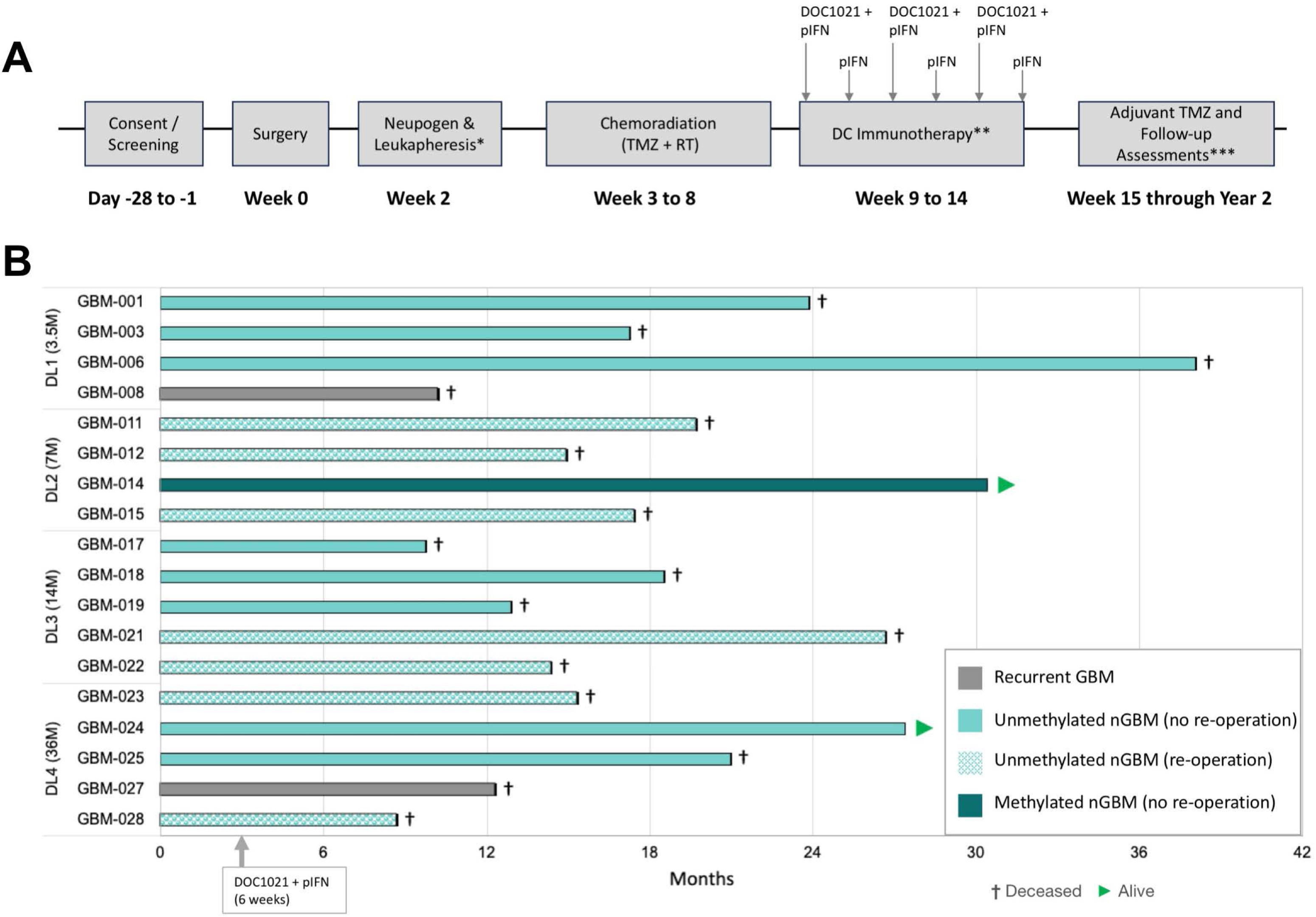
Study schema and swimmer plot. **A.** Timeline of study related activities. *Neupogen was administered starting 5 days before leukapheresis; **DC immunotherapy consisted of 3 doses of DOC1021 via perinodal injection (14 days apart, with weekly pIFN); ***Adjuvant TMZ consisted of 5 consecutive days of TMZ every 28-day cycle, for 6-12 cycles. Brain MRIs were performed as per SOC and patients were followed for survival. NOTE: Recurrent GBM patients could have followed the same treatment protocol/timing as for newly diagnosed GBM (shown here) or had an abbreviated treatment schedule, with DC immunotherapy beginning sooner at the discretion of the neuro-oncology treatment team. **B.** Swimmer plot showing survival of nGBM and recurrent GBM patients starting from time of surgery. DOC1021 + peg-IFN administered over 6 weeks after completion of chemoradiation, prior to initiating adjuvant TMZ. Teal bars- nGBM patients, grey bars- recurrent GBM patients, dark teal- MGMT methylated patient, lighter teal-MGMT unmethylated patients, hash marked bars- re-operated within 6 months of vaccination, solid bars- not re-operated. DC: dendritic cell; MRI: magnetic resonance imaging; pIFN: pegylated interferon alpha 2a; RT: radiotherapy; TMZ: temozolomide

Patient characteristics are shown in **Table 1** (removed to comply with medRxiv policy). For the 16 nGBM patients, median age was 61 years; 50% were female; ECOG at baseline was 0 in 38%, 1 in 56% and 2 in 6%. All tumors were IDH-wt. This study population had a worse prognosis than most comparison study populations given that only 1 of 18 subjects were classified as MGMT promoter methylated. Further, significant residual tumor tissue after surgery was present in 4 patients (subtotal or partial resection) and progression prior to vaccine administration was not exclusionary and occurred in 5 patients (2 with subtotal and 3 with near-total resection). After DOC1021 administration, re-operation for suspected progression occurred within 6 months in 7 nGBM patients. In addition to adjuvant TMZ, 9 patients (GBM-008, 011, 012, 015, 017, 018, 024, 025 and 028) received tumor-treating fields for varying durations. After progression, additional treatments included bevacizumab, supplemental radiation and various salvage chemotherapies including CCNU.

### Safety

Adverse events (AEs) occurring from the time of first vaccination through 30 days after the final DOC1021 vaccination that were deemed possibly, probably, or definitely related to the vaccine product are listed in **Table 2** with all AEs within this time-period listed in **Table S1**. There were no dose-limiting toxicities, serious adverse events or AEs greater than grade 2 that were deemed related (possibly, probably, or definitely) to the product. There were no notable differences in AEs between dose levels. The most common AEs related to the vaccine were low-grade (grade 1-2) and transient flu-like symptoms (including fatigue, headache, arthralgia, chills, nausea, vomiting),, and injection site reactions (including neck pain, pruritus, and urticaria) managed with standard analgesics, anti-histamines and/or topical medications. Two patients experienced transient > grade 2 cytopenias (neutropenia and leukopenia grade 3 in one and neutropenia grade 4 and thrombocytopenia grade 3 in the other) that are known side effects of both peg-IFN and temozolomide chemotherapy.

**Table 2.** Treatment-Emergent Related AEs.

| **System Organ Class**<br>**Preferred Term** | **Grade 1** | **Grade 2** | **Grade**<br>**≥3** | **Total** |
| --- | --- | --- | --- | --- |
| **Blood and lymphatic system disorders** | **2** | **--** | **--** | **2** |
| *Eosinophilia* | 1 | -- | -- | 1 |
| *Lymph node pain* | 1 | -- | -- | 1 |
| **Ear and labyrinth disorders** | **--** | **1** | **--** | **1** |
| *Vestibular Disorder* | -- | 1 | -- | 1 |
| **Gastrointestinal disorders** | **9** | **2** | **--** | **11** |
| *Abdominal pain* | 1 | -- | -- | 1 |
| *Nausea* | 6 | 2 | -- | 8 |
| *Vomiting* | 2 | -- | -- | 2 |
| **General disorders and administration site conditions** | **18** | **8** | **--** | **26** |
| *Chills* | 2 | 1 | -- | 3 |
| *Fatigue* | 6 | 7 | -- | 13 |
| *Flu-like symptoms* | 3 | -- | -- | 3 |
| *Injection site reaction* | 5 | -- | -- | 5 |
| *Localized edema* | 1 | -- | -- | 1 |
| *Non-cardiac chest pain* | 1 | -- | -- | 1 |
| **Injury, poisoning and procedural complications** | **1** | **--** | **--** | **1** |
| *Bruising* | 1 | -- | -- | 1 |
| **Investigations** | **1** | **--** | **--** | **1** |
| *Platelet count decreased* | 1 | -- | -- | 1 |
| **Musculoskeletal and connective tissue disorders** | **16** | **--** | **--** | **16** |
| *Arthralgia* | 2 | -- | -- | 2 |
| *Back Pain* | 1 | -- | -- | 1 |
| *Generalized muscle weakness* | 1 | -- | -- | 1 |
| *Myalgia* | 2 | -- | -- | 2 |
| *Neck pain* | 10 | -- | -- | 10 |
| **Nervous system disorders** | **7** | **2** | **--** | **9** |
| *Dysgeusia* | 2 | -- | -- | 2 |
| *Headache* | 5 | 1 | -- | 6 |
| *Lethargy* | -- | 1 | -- | 1 |
| **Psychiatric disorders** | **--** | **1** | **--** | **1** |
| *Confusion* | -- | 1 | -- | 1 |
| **Reproductive system and breast disorders** | **1** | **--** | **--** | **1** |
| *Irregular menstruation* | 1 | -- | -- | 1 |
| **Skin and subcutaneous tissue disorders** | **12** | **3** | **--** | **15** |
| *Pruritus* | 7 | -- | -- | 7 |
| *Rash maculo-papular* | 2 | -- | -- | 2 |
| *Urticaria* | 3 | 3 | -- | 6 |
| **Grand Total** | **67** | **17** | **0** | **84** |

### Efficacy

For nGBM patients from time of surgery, 88% remained alive at 12 months and after median follow-up of 30.4 months, two patients remained alive at the date of data cutoff (**Figure 1B**). Kaplan Meier survival analysis (**Figure S1**) revealed a median OS of 18.5 months for nGBM patients. The two recurrent patients survived 10.2 and 12.3 months (**Figure 1B**). Cause of death was related to GBM progression or recurrence in all patients except GBM-001, who died of unknown cause 7 months after entering hospice following COVID-19 and multiple falls without further GBM follow up, and GBM-003 who died by medically assisted suicide following progression.

Magnetic resonance images for the two longest-lived patients, GBM-006 and GBM-014, demonstrated increased T1 contrast-enhancing signal appearing 1-2 months after vaccination but gradually resolving over subsequent months with complete resolution by month 14-18 (**Figure 2**). Patient GBM-006 survived for 38.1 months, eventually dying from presumed recurrence in a new location. Patient GBM-014 remains alive without recurrence at 30.4 months. Patient GBM-024 also remains alive with good performance status at 27.7 months. These patients did not receive re-operation or bevacizumab. In contrast, 7 patients that developed increased T1-signal 6-23 weeks after first DOC1021 administration were treated with re-operation despite no significant clinical decline and an ECOG maintained at 0 or 1. Survival among re-operated patients shows a substantial trend toward worse outcome than those who did not receive re-operation though small numbers preclude statistical comparison (**Figure S1**). Baseline characteristics of patients who received reoperation versus those who did not are shown in **Table S2**.

**Figure 2.**
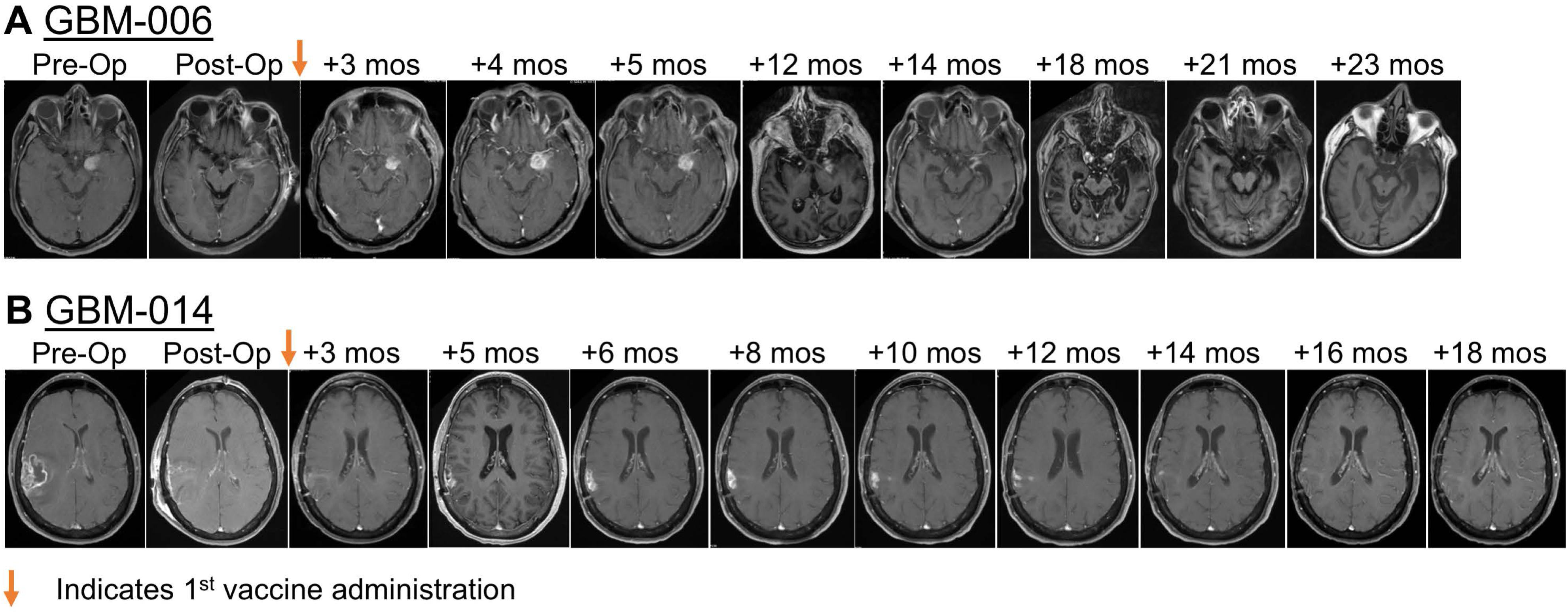
Longitudinal radiologic assessment of long-lived patients. Serial T1-weighted, contrast-enhanced axial MRI images **A.** Newly-diagnosed MGMT unmethylated patient GBM-006 longitudinal imaging (received 3.5 x 10^6^ dendritic cells, 38.1 mos survival). **B.** Newly-diagnosed MGMT methylated patient GBM-014 longitudinal imaging (received 7 x 10^6^ dendritic cells, > 30.4 mos survival). These patients did not receive re-operation or bevacizumab.

### Post-vaccination Analysis of the Tumor Immune Microenvironment

Tumor samples available from 3 patients who underwent early postvaccination reoperation for enhancing MRI signal provided the opportunity to perform spatial transcriptomic analysis on pre- and post-vaccination FFPE tumor samples. Using a predefined transcriptomic reference signature (**Figure S2A**)[35], normal brain, immune cell, and cancer cell subtypes were defined and plotted for all six samples (**Figure S2B**). In concordance with the pathology reports of these patients, only one of three patients (GBM-011) exhibited pathologic relapse, i.e. presence of tumor cells in the re-operation specimen (**Figures S2B/C**). The absence of tumor cells in GBM-012 and GBM-015 coupled with substantially upregulated T-cell infiltration (**Figures S2C/S3A**) in these patients suggested clinical pseudo-progression rather than true progression. Increased numbers of T-cells in the post-vaccination specimens was confirmed in patients GBM-012 and GBM-015 by cell2location deconvolution (**Figure S3B**) as was substantially increased myeloid cell infiltration among all three patients (**Figures S3A/B**).

Next, *immune triad analysis* was performed, mapping the colocalization of hematopoietic-derived migratory microglia[36] with inflammatory, activated CD4^+^ T-cells and activated CD8^+^ cells expressing critical memory markers, CD161, CD127, and CD62L (triad gene signature defined in **Figure 3A**). The incidence of such triads was significantly increased in post-vaccination samples of all three patients analyzed (**Figures 3B/C**). Interestingly, the presence of regulatory T cells (T-regs) declined in the two patients who had not relapsed at the time of re-operation but remained constant in the patient who did exhibit pathologic relapse (**Figures 3A/C**). Analyzing the proximity/density of immune triads, both to each other and to T-regs, demonstrated that triads of inflammatory CD4^+^ cells/memory CD8^+^ cells/migratory microglia were significantly more dense and tightly clustered in post-vaccination than in pre-vaccination specimens (**Figure 3D**). Moreover, post-vaccination immune triads appeared to actively exclude T-regs which, when present, were much farther afield than in all pre-vaccination samples **(Figure 3D**). These data suggest the occurrence of active post-vaccination immune processes mediated by migratory, microglia-like cells of hematopoietic origin acting in concert with effector memory CD8^+^ cells and inflammatory CD4^+^ cells in all three of the patients analyzed.

**Figure 3.**
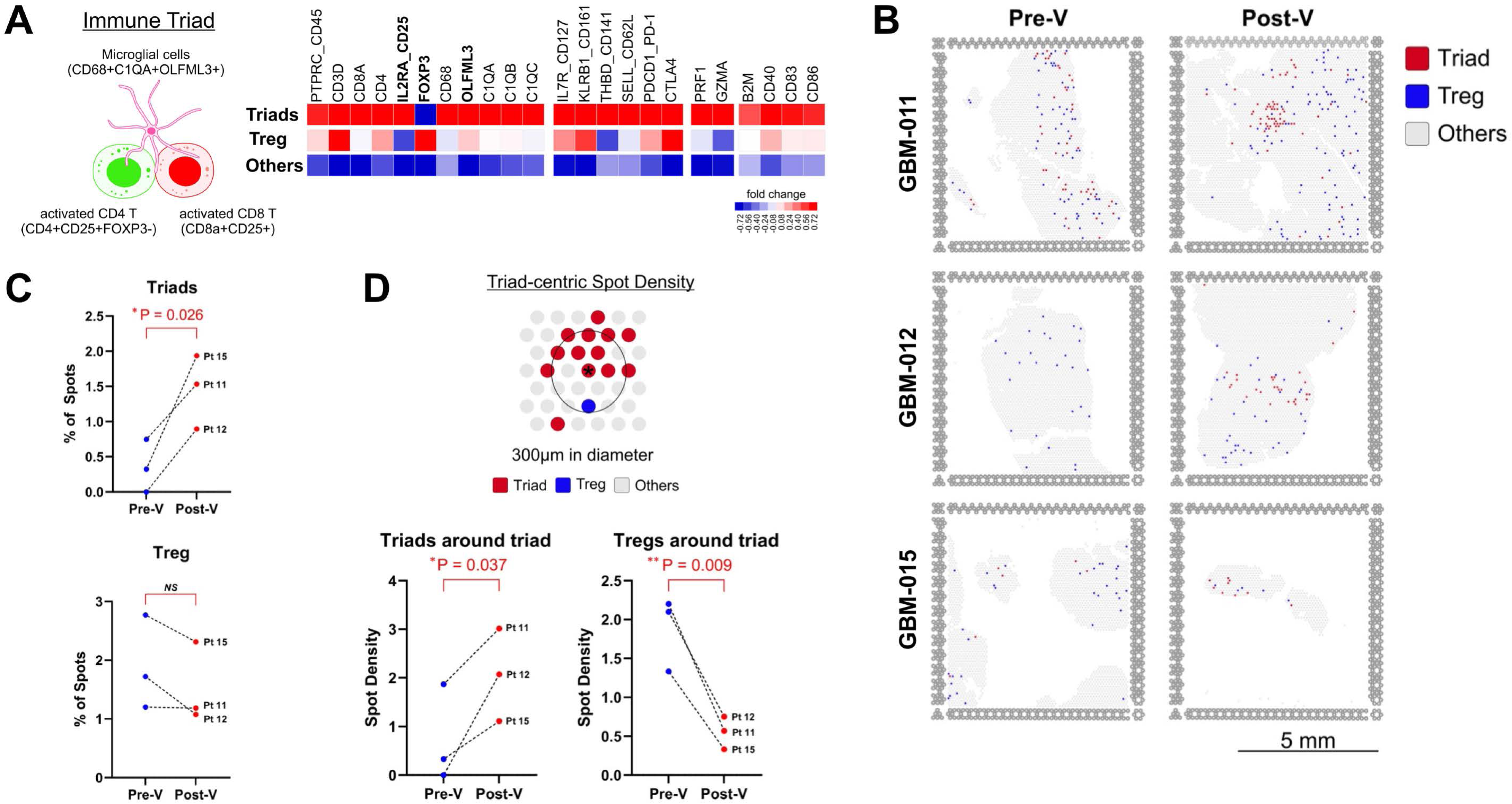
Spatial characterization of immune cell triads in glioblastoma pre/post-vaccination. Spatial transcriptomic analysis of pre- and post-vaccination FFPE tumor samples from pre-vaccination (pre-V, time of initial resection) and post-vaccination (post-V, at 14 weeks (GBM-011), 23 weeks (GBM-012) and 6 weeks (GBM-015) after initial DOC1021 vaccination. **A.** Transcriptional definition and schematic representation of an immune cell triad, composed of activated CD4⁺ T-cells (CD4⁺CD25⁺Foxp3⁻), activated CD8⁺ T-cells (CD8a⁺CD25⁺), and microglial cells (CD68⁺C1qa⁺OLFML3⁺). The accompanying heatmap shows the relative gene expression (log2 fold-change) across triad, Treg, and other cell types for selected immune and microglial markers in spatial transcriptomic spots. Key genes defining triads (e.g., IL2RA, Foxp3, CD25, OLFML3, C1qa) are highlighted. **B.** Spatial maps of triad (red), treg (blue), and other (gray) cell-type-enriched spots in three patients analyzed before (pre-V) and after (post-V) vaccination. Red spots indicate triad-enriched areas, showing increased distribution and clustering post-vaccination. **C.** Postvaccination changes in triad- and treg-enriched spots in each patient. **D.** Immune triad-centric spot density analysis within a 300 µm radius. Following vaccination, treg density significantly decreased, while the density of immune triads significantly increased around triad-enriched regions, suggesting enhanced spatial organization and coordination of the immune response. For all panels, *p < 0.05, **p < 0.01 by Student’s two-tailed t-test.

### Immunophenotypic Analysis of Postvaccination Peripheral Blood

Peripheral blood was collected from patients at baseline and two weeks after administration of the final vaccine product. Analysis of these time points by flow cytometry (**Figure 4, S4 and S5-S32**) then identified a number of consistent and significant alterations to peripheral blood immune cell populations following the vaccination regimen. Particularly, we observed significant postvaccination expansions of both CD4^+^ (CD45RA^neg^CD45RO^+^CD62L^+^CCR7^+^), and CD8^+^ (CD45RA^neg^CD45RO^+^CD62L^+^ CCR7^+^) central memory T cells (T_CM_) (**Figures 4A and S4A/B**) with additional significant expansion of CD4^+^CD161^+^ (**Figures 4B and S4C**), a critical cell population associated with complete responses to immunotherapy and good outcomes in HPV^+^ tumors.[37, 38] CD8^+^CD127^+^ long-lived memory precursor cells (MPECs[39–41]) were substantially enhanced in post-vaccination peripheral circulation, (**Figures 4A-C and S4D**) suggesting the development of durable immunologic memory.

**Figure 4.**
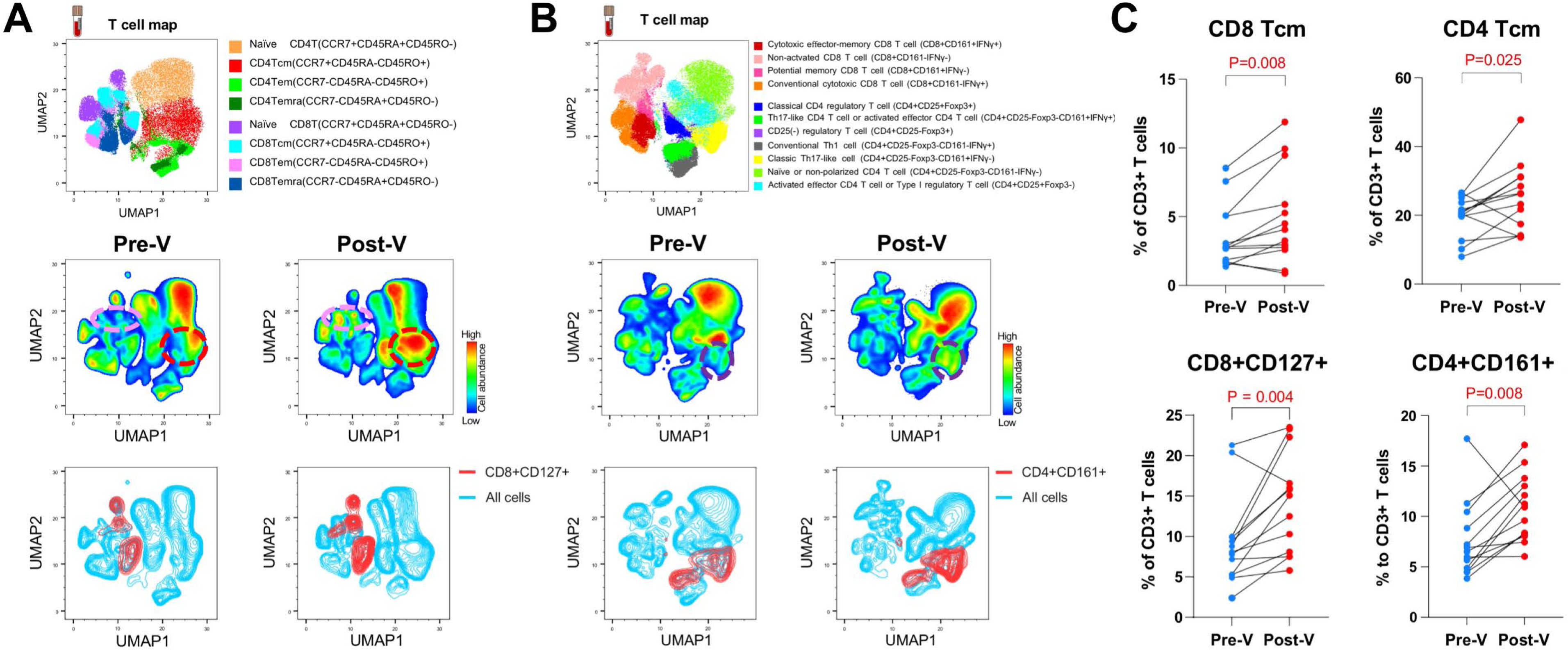
Immunophenotypic alterations in peripheral blood following DOC1021 vaccination. Peripheral blood collected from patients at baseline and again at day 106-108 of the experimental regimen were characterized by flow cytometry. **A/B.** Phenotypic and functional changes in CD4⁺ and CD8⁺ T-cell memory (**A**) and functional (**B**) subsets. UMAP clustering of CD3⁺ T-cells was annotated by conventional memory subsets based on CD45RA, CD45RO, and CCR7 expression, identifying naïve, central memory (Tcm), effector memory (Tem), and terminally differentiated effector (Temra) populations. UMAP density plots pre- and post-vaccination highlighted expansion of CD8^+^ T_CM_ (pink dotted circle), CD4^+^ T_CM_ (red dotted circle), and CD4^+^CD161^+^ (purple dotted circle) T-cell subsets after vaccination. Upregulation of CD127^+^ MPECs is shown separately (blue/pink UMAP plots), highlighting the upregulation of CD127 across multiple traditional phenotypic populations. **C.** Paired comparison of selected T-cell subsets as a percentage of total CD3⁺ T-cells before and after vaccination showing significant increases in CD4⁺ Tcm, CD8⁺ Tcm, CD8^+^CD127^+^ MPECs and CD4⁺CD161⁺ effector T-cells. For each panel, *p<0.05, **p<0.01, ***p<0.005, by paired Student’s two-tailed t test.

## Discussion

Successful application of immunotherapy to GBM faces significant hurdles including the immunosuppressive tumor microenvironment, tumor antigenic heterogeneity, and development of therapeutic resistance. Nevertheless, the infiltrative nature of GBM and risk of toxicity to normal brain are substantial obstacles to standard cancer treatment approaches that incentivize continued development of immune-based therapies. In the immunotherapeutic approach described here, multiple innate and adaptive signals characteristic of viral infection are combined in a way that fosters generation of immunologic memory. The loading of homologous antigenic determinants on dendritic cell MHC class I and II serves as a powerful virus-associated PAMP that influences development of T_H_1 polarizing immune responses through its ability to induce qualitative differentiation of DC populations [20, 23, 24, 26, 29, 30, 42–44]. Monocyte-derived DC pre-conditioned by exposure to this antigen-based PAMP differentiate into cDC1-like cells that, in *in vitro* studies, generate ∼log-fold greater numbers of CD8^+^IFN-γ^+^ cytolytic effectors than DC loaded with mRNA or tumor lysate only.[24] This enhanced immune response includes development of CD161^+^ effectors possessing critical properties that include a propensity for serial killing, inherent tissue homing capabilities, and a substantial degree of resistance to functional exhaustion [27, 30, 45–49]. The sensing mechanism through which homologous antigenic epitopes are detected is integrated within broader innate inflammatory cascades which, together with nucleic acid pattern recognition receptors (PRRs), comprise a critical component of anti-viral signaling that informs adaptive immune T_H_ polarization.[20–23]

In the present phase I clinical trial, DOC1021 (dubodencel), a homologous, doubly-loaded DC preparation, was combined with type I interferon adjuvantation of the patient to mimic characteristics of physiologic viral infection. The clinical protocol also stipulated delivery of the vaccine product in the proximity of the deep cervical lymph node chains, the physiologic sites of CNS immune cell priming.[50] Adaptive immune T-cells are programmed during priming to express cell surface addressin molecules, integrins and chemokine receptors that permit homing back to those sites and extravasation through binding to compartment-specific receptors on vascular endothelial cells. In the specific instance of the CNS, T-cells primed in the deep cervical nodes are programmed to express the α_4_β_1_ integrin VLA-4 which binds to inducibly-expressed VCAM-1 on the inflamed CNS vascular endothelium.[51–54] In addition to enhancing T-cell killing of tumor [55, 56], type I interferon may also assist in upregulating endothelial surface expression of VCAM-1 thereby improving T-cell localization to the CNS [57]. Accordingly, vaccines administered intra-dermally or intravenously and/or without appropriate adjuvantation might not induce optimal trafficking of reactive T-cells to the CNS.[52]

The DOC1021 regimen was found to be feasible and safe without dose limiting or apparent dose-related toxicities in combination with SOC for GBM. Analysis of the post-vaccination tumor microenvironment after DOC1021 in three patients demonstrated that T cells successfully crossed the blood brain barrier and homed to the tumor. Post-vaccination immune microenvironments were notable for the presence of immune triads composed of activated CD4^+^ and CD8^+^ T-cells and migratory microglial cells that appeared to actively exclude tregs. Postvaccination alterations to peripheral blood T-cell immunophenotypes were also consistent with mechanism as defined in preclinical studies including increased levels of CD4^+^ and CD8^+^ T_CM_, CD8^+^CD127^+^ long-lived memory precursor effector cells (MPECs [39–41]) and CD161^+^ effector CD4^+^ T-cells. CD4^+^CD161^+^ T-cells are particularly interesting given the prominent associations of these with complete response to immunotherapy as well as good outcomes in certain tumor types.[37, 38] In sum, the data suggested the hypothesis that DOC1021 facilitates development of effector memory CD8^+^ T-cells that may ingress and reside locally for months, remaining active through local antigen presentation by inflammatory myeloid cells.

Imaging is an imprecise tool for evaluation of GBM treatment effect, especially so for immunotherapies due to edema and inflammation that cannot be easily differentiated from true tumor progression by MRI alone.[58–60] The gradual resolution of increased T1-signal observed in some patients in this study and the pathologic and immune microenvironmental changes in samples from re-operated patients suggest that inflammation (i.e. pseudo-progression) is commonly observed after DOC1021 administration for GBM. Re-operation within 6 months after DOC1021 vaccination appeared to negatively impact survival, perhaps due to removal of an immunostimulatory microenvironment and/or administration of immunoinhibitory dexamethasone at the time of re-operation. While small sample size precluded definitive conclusion, survival was encouraging considering the poor prognosis of the study population. The study permitted enrollment of only IDH-wt patients without an upper age limit, allowed any degree of resection, did not exclude patients who exhibited progression prior to vaccine administration, and was comprised almost exclusively MGMT promoter unmethylated (15/16, 94%) disease. This is a population expected to exhibit substantially worse outcomes than contemporaneous studies with stringent inclusion criteria or historic studies that did not exclude IDH mutant disease. [61] The 12 month OS of 88% (or 87% among the 15 MGMT unmethylated patients) and median OS of 18.5 months compare favorably to MGMT unmethylated cohorts reported by Hegi et. al. 2005 (12 month OS 58.3%, median OS 12.7 months) [7], Stupp 2017 (12 month OS 63.8%, median OS 14.7 months) [4], and Omuro 2023 (12 month OS 61.2%, median OS 14.9 months). [19]

Spatial identification and characterization of inflammatory immune cell triads in the post-vaccination tumor bed was a significant finding of this study. Recent literature suggests that the presence of triad complexes comprising local antigen presenting cells, CD8^+^ T-cells, and inflammatory CD4^+^ helper T-cells are a requirement for the elimination of solid tumor in the context of immunotherapy.[62] Consistent with this critical theme, reoperation explants taken from three patients early (1-4 months) in the postvaccination period showed substantial upregulation of triad complexes comprised of effector memory CD8^+^ cells, inflammatory helper CD4^+^ T-cells, and microglia derived from migratory hematopoietic precursors but not yolk-sac derived resident populations (e.g. **Figure 3**). Interestingly we observed not only an upregulation of total triad complexes in all three patients examined, but also increased spatial clustering of triads as well as exclusion of regulatory T-cells from the vicinity of triad clusters.

In summary, we describe a novel cell-based vaccination methodology for the adjuvant treatment of GBM that leverages recent advances in the understanding of dendritic cell biology and T-cell homing. This treatment regimen was shown to be both safe and feasibly integrated within existing standard of care. While conclusions regarding OS must be interpreted with caution in open label trials of limited size, results suggest enhanced survival, particularly among participants that did not receive reoperation. Clinical findings were reinforced by characterization of increased memory T-cell populations in the postvaccination peripheral blood and upregulation of inflammatory immune cell triad complexes in the postvaccination tumor bed. These data support additional investigation, including the initiation of a Phase II randomized, controlled trial for nGBM (NCT06805305).

## Disclosure

WKD, VK, JH, MMH and JFG declare an ownership stake in Diakonos Research, Ltd. WKD, VK, JH, DO, IB, EHS, EMD and LKA declare financial compensation and in some cases equity options from Diakonos Oncology Corporation. WKD declares an unrelated financial relationship with APAC Biotech, Pvt, Ltd from 2015 to 2020. LKA declares unrelated stock ownership in Candel Therapeutics. SHH declares unrelated equity options and financial compensation from CNS Pharmaceuticals. J-JZ declares unrelated industrial grant support from Novocure, Plus Therapeutics, Five Prime Therapeutics, ABM Therapeutics, and Boston Biomedical of Dainippon Sumimoto Pharma as well as financial compensation from Novocure. NT declares unrelated equity options and financial compensation from BrainDynamics and financial compensation from Nervonik. All remaining authors declare no financial conflicts of interest.

## Author Contributions

JFG was the study sponsor, designed and led clinical studies, and analyzed data. CC led clinical studies and provided patient care. SA provided patient care and supported clinical studies. AG supported clinical studies. CM led clinical studies and provided patient care. CJB led clinical studies and provided patient care. AV supported clinical studies and provided patient care. JI analyzed data. MZ supported clinical studies and analyzed data. LC supported clinical studies. LK supported clinical studies. EHS supported clinical studies. MV supported clinical studies. BAB provided biostatistical support and analyzed data. ATrivedi manufactured vaccines and supported clinical studies. WL manufactured vaccines and supported clinical studies. NM manufactured vaccines. CJH manufactured vaccines and supported clinical studies. KJE performed experiments and analyzed data. NB performed experiments. JV-P performed experiments. DO-S performed experiments. SBA performed experiments. VRodriguez performed experiments. DCK analyzed data. DO supported clinical studies. IB supported clinical studies, analyzed data and wrote the paper. JH supported clinical studies. MMH analyzed data. EMD analyzed data and wrote the paper. LKA analyzed data, supported clinical studies, and wrote the paper. SHH led clinical studies and provided patient care. J-JZ led clinical studies and provided patient care. RCZ provided patient care. YB provided patient care. SWK provided biostatistical support and analyzed data. H-JJ provided biostatistical support and analyzed data. HSL provided biostatistical support and analyzed data. TE analyzed data. RG analyzed data. YE-L served as clinical site PI, led clinical studies, and provided patient care. NT led clinical studies and provided patient care. ATurtz served as clinical site PI, designed and led clinical studies, and provided patient care. VK manufactured vaccines, supported clinical studies, performed experiments, and analyzed data. WKD designed and supported clinical studies, analyzed data, and wrote the paper.

## Data Availability

The Visium spatial transcriptomic data that support the findings of this study are available in the Gene Expression Omnibus (GEO) at http://ncbi.nlm.nih.gov/geo, accession number GSE286477. Additional data are available upon reasonable request to the senior author.

## Acknowledgements

The study authors are indebted to Liz and Jay Scott and Lisa Towry of Alex’s Lemonade Stand Foundation for their many years of support. The authors are similarly grateful for the unwavering support of Tom, Dan, Mike and countless others affiliated with Diakonos Oncology Corporation. This work is dedicated to the memories of friends and collaborators Dr. Jonathan M. Levitt and Dr. Robert J. Arceci, both of whom passed away unexpectedly before trial enrollment could begin, and Mr. Adam Chachkes who courageously fought GBM before succumbing in 2023. This study was supported in part by a grant from Cancer Cures 4 Kids (to WKD) with additional support from Diakonos Oncology Corporation. This study was further supported in part by the Baylor College of Medicine flow cytometry core facility under the academic leadership of Dr. Christine Beeton and the expert assistance of Mr. Joel M. Sederstrom. This study was additionally supported in part by the Genomic and RNA Profiling Core at Baylor College of Medicine with funding from NIH S10 grant 1S10OD023469, NCI grant P30CA125123, CPRIT grant RP250580, and outstanding technical support from M. Sayeed Sayeeduddin, Shahida Salar, and Zahida Sayeeduddin (histology); Myoung Kwon and Maciej Zalazaoski, Ph.D. (immunohistochemistry and imaging), and Marcus Cabrera (tissue and data collection). Lastly, this work was additionally supported in part by NIH NCI grant R37 CA248478, US Department of Defense Impact Award W81XWH-22-1-0657, NIH NIAID grant R21 AI159379, and a Helis Medical Research Foundation grant (to HSL).

## Supplemental Figure legends

**Figure S1. Kaplan-Meier survival curves of nGBM patients. A.** OS from time of surgery for 16 nGBM patients. Median OS 18.5 months (black dotted line). **B.** Comparison of OS for patients that did (n=7) or did not (n=9) receive re-operation within 6 months after DOC1021 vaccination. Median OS 23.8 months for no re-operation vs 15.3 months for re-operation group (p=0.06 by log-rank).

**Figure S2. Spatial transcriptomic analyses of the pre- and post-vaccination tumor bed.** Spatial transcriptomic analysis of pre- and post-vaccination FFPE tumor samples was performed as described. **A.** Predefined transcriptomic reference signature of brain parenchyma cell types was derived from Wang etl al.[35] **B.** Composite Umap plot of all cell clusters identified in three paired pre- and post-vaccination FFPE samples. **C.** Individual Umap plot of each patient’s pre- and post-vaccination FFPE sample with cancer cells identified post-vaccination in only one of three patients in concordance with postsurgical pathology reports.

**Figure S3. Quantitative analysis of select cell populations in the pre- and post-vaccination tumor bed. A.** Post-vaccination changes to T-cell, myeloid cell, neuronal cell, oligodendrocyte, pericyte-endothelial cell, and fibroblast cell populations. *p < 0.05 as determined by Student’s two-tailed t-test. **B.** Cell2location deconvolution was employed to determine the total proportion of T-cells and myeloid cells per spot with statistical significance determined by Student’s two-tailed t-test and *p* values shown.

**Figure S4. Enumeration of immunophenotypic alterations to peripheral blood immune cell populations following DOC1021 vaccination.** Peripheral blood collected from patients at baseline and again at day 106-108 of the experimental regimen were characterized by flow cytometry. Raw percentage changes in T-cell populations of individual patients for whom data were available (of total CD4^+^ or CD8^+^ cells as indicated) are shown in black with average percent change shown in red. **A.** CD3^+^CD4^+^CD45RA^neg^CD45RO^+^ CD62L^+^CCR7^+^ TCM, expansion from pre-vaccination mean 33.7% to post-vaccination mean 46.2%. **B.** CD3^+^CD8^+^CD45RA^neg^CD45RO^+^ CD62L^+^CCR7^+^ TCM, expansion from pre-vaccination mean 17.5% to post-vaccination mean 23.0%. **C.** CD3^+^CD4^+^CD161^+^ effector cells, expansion from pre-vaccination mean 16.0% to post-vaccination mean 21.7%. **C.** CD3^+^CD8^+^ CD127^+^ MPECs, expansion from pre-vaccination mean 34.6% to post-vaccination mean 49.9%. For each panel, ***p<0.005, ****p<0.001 by paired Student’s two-tailed t test.

**Figure S5. Flow cytometry panel 1 gating and analysis strategy using healthy donor PBMC control cells.** Shown: Zombie UV viability dye. CD3-APC. CD4-PE-Cy5. CD8-BUV737. CD161-PE. CD25-AF488. IFN-γ-BV711. Granzyme B-APC-Cy7. Foxp3-BV421.

**Figure S6. Flow cytometry and analysis panel 1 for patient GBM-011.**

**Figure S7. Flow cytometry and analysis panel 1 for patient GBM-012.**

**Figure S8. Flow cytometry and analysis panel 1 for patient GBM-014.**

**Figure S9. Flow cytometry and analysis panel 1 for patient GBM-015.**

**Figure S10. Flow cytometry and analysis panel 1 for patient GBM-017.**

**Figure S11. Flow cytometry and analysis panel 1 for patient GBM-018.**

**Figure S12. Flow cytometry and analysis panel 1 for patient GBM-019.**

**Figure S13. Flow cytometry and analysis panel 1 for patient GBM-021.**

**Figure S14. Flow cytometry and analysis panel 1 for patient GBM-022.**

**Figure S15. Flow cytometry and analysis panel 1 for patient GBM-023.**

**Figure S16. Flow cytometry and analysis panel 1 for patient GBM-025.**

**Figure S17. Flow cytometry and analysis panel 1 for patient GBM-027.**

**Figure S18. Flow cytometry and analysis panel 1 for patient GBM-028.**

**Figure S19. Flow cytometry panel 2 gating and analysis strategy using healthy donor PBMC control cells.** Shown: Zombie UV viability dye. CD3-APC. CD4-PE-Cy5. CD8-BUV737. CCR7-PE. IFN-γ-BV711. CD45RO-BV421. CD45RA-AF488. CD127-APC-Cy7. CD62L-RB780.

**Figure S20. Flow cytometry and analysis panel 2 for patient 0011.**

**Figure S21. Flow cytometry and analysis panel 2 for patient 0012.**

**Figure S22. Flow cytometry and analysis panel 2 for patient 0014.**

**Figure S23. Flow cytometry and analysis panel 2 for patient 0015.**

**Figure S24. Flow cytometry and analysis panel 2 for patient 0017.**

**Figure S25. Flow cytometry and analysis panel 2 for patient 0018.**

**Figure S26. Flow cytometry and analysis panel 2 for patient 0019.**

**Figure S27. Flow cytometry and analysis panel 2 for patient 0021.**

**Figure S28. Flow cytometry and analysis panel 2 for patient 0022.**

**Figure S29. Flow cytometry and analysis panel 2 for patient 0023.**

**Figure S30. Flow cytometry and analysis panel 2 for patient 0025.**

**Figure S31. Flow cytometry and analysis panel 2 for patient 0027.**

**Figure S32. Flow cytometry and analysis panel 2 for patient 0028.**

## Supplemental Methods

### Flow Cytometry

Cryopreserved PBMC were thawed and rested overnight in complete RPMI 1640 culture medium (Gibco) supplemented with 10% human serum and antimycotic-antibiotic (cRPMI, ThermoFisher Scientific, Waltham, MA) at 37°C. Rested PBMC were treated with ionomycin and phorbol 12-myristate 13-acetate and incubated for 4 hours at 37° C in cRPMI. Cells were then washed twice with Dulbecco’s phosphate buffered saline (DPBS, ThermoFisher), resuspended in Zombie UV™ Fixable Viability Dye (BioLegend, San Diego, CA), and incubated at room temperature for 15 minutes. Cells were washed twice with FACS buffer (2% fetal calf serum in DPBS) before surface staining with antibody suspended in FACS buffer for 30 minutes at 4° C. Afterwards, cells were washed twice with FACS buffer and intracellular staining was performed with Foxp3/Transcription Factor Fixation/Permeabilization Kit (ThermoFisher) according to manufacturer’s instructions with the exception that intracellular staining was performed for 1 hour at room temperature instead of 30 minutes. After the final wash step, cells were acquired at ≤ 2000 events/second on a FACSymphony A5 flow cytometer (BD, Franklin Lakes, NJ). Data analysis was performed using FlowJo software v10.10 (BD).

### Flow Antibody Reagents

CD3-APC (BioLegend Cat #423108; RRID: AB_1937212), CD4-PE/Cy5 (BioLegend Cat #344654; RRID: AB_2810545), CD8a-BUV737 (eBioscience Cat #367-0088-41; RRID: AB_2895930), IFN-γ-BV711, BioLegend Cat #502540; RRID: AB_2563506), CD161-PE (BioLegend Cat #339938; RRID: AB_2564141), CD25-AF488 (BioLegend Cat # 356116; RRID: AB_2562166), granzyme B-APC/Cy7 (BioLegend Cat # 372227; RRID: AB_2936711), Foxp3-BV421 (BioLegend Cat #320124; RRID: AB_2565972), CD45RA-AF488 (BioLegend Cat #304114; RRID: AB_528816), CD45RO-PacBlue (BioLegend Cat #304216; RRID: AB_493659), CCR7-PE (BioLegend Cat #353203; RRID: AB_10916391), CD127-APC/Cy7 (BioLegend Cat #351347; RRID: AB_2629571), CD62L-RB780 (BD Biosciences Cat #569211; RRID: AB_3668758),

Mouse IgG_1_κ Isotype-AF488 (BioLegend Cat #400132; RRID: AB_2890263), Mouse IgG_1_ κ Isotype-BV711 (BioLegend Cat #400167; RRID: AB_11218607), Mouse IgG_1_ κ Isotype-PE (BioLegend Cat #400112; RRID: AB_2847829), Mouse IgG_1_ κ Isotype-RB780 (BD Biosciences Cat #568532; RRID: AB_3668760), Mouse IgG_1_ κ Isotype-APC/Cy7 (BioLegend Cat #400128; RRID: AB_2892538), Mouse IgG_2a_ κ Isotype-Pac-Blue (BioLegend Cat #400235; RRID: AB_493447), Mouse IgG_2a_ κ Isotype-BV421 (BioLegend Cat #400259; RRID: AB_10895919).

---

**Table S1.** Treatment-Emergent Related and Unrelated AEs.

| System Organ Class<br>*Preferred Term* | Grade 1 | Grade 2 | Grade 3 | Grade 4 | Total |
| --- | --- | --- | --- | --- | --- |
| **Cohort 1** | **40** | **23** | **2** | **--** | **65** |
| **Ear and labyrinth disorders** | **--** | **1** | **--** | **--** | **1** |
| *Vestibular Disorder* | *--* | *1* | *--* | *--* | *1* |
| **Gastrointestinal disorders** | **8** | **1** | **--** | **--** | **9** |
| *Nausea* | *6* | *1* | *--* | *--* | *7* |
| *Vomiting* | *2* | *--* | *--* | *--* | *2* |
| **General disorders and administration site conditions** | **5** | **7** | **--** | **--** | **12** |
| *Chills* | *2* | *1* | *--* | *--* | *3* |
| *Fatigue* | *2* | *6* | *--* | *--* | *8* |
| *Flu-like symptoms* | *1* | *--* | *--* | *--* | *1* |
| **Injury, poisoning and procedural complications** | **2** | **--** | **--** | **--** | **2** |
| *Fall* | *2* | *--* | *--* | *--* | *2* |
| **Investigations** | **9** | **5** | **2** | **--** | **16** |
| *Neutrophil count decreased* | *3* | *3* | *1* | *--* | *7* |
| *Platelet count decreased* | *3* | *--* | *--* | *--* | *3* |
| *White blood cell decreased* | *3* | *2* | *1* | *--* | *6* |
| **Metabolism and nutrition disorders** | **--** | **1** | **--** | **--** | **1** |
| *Anorexia* | *--* | *1* | *--* | *--* | *1* |
| **Musculoskeletal and connective tissue disorders** | **3** | **--** | **--** | **--** | **3** |
| *Neck pain* | *3* | *--* | *--* | *--* | *3* |
| **Nervous system disorders** | **8** | **3** | **--** | **--** | **11** |
| *Ataxia* | *--* | *1* | *--* | *--* | *1* |
| *Edema cerebral* | *1* | *--* | *--* | *--* | *1* |
| *Headache* | *4* | *1* | *--* | *--* | *5* |
| *Hydrocephalus* | *--* | *1* | *--* | *--* | *1* |
| *Memory impairment* | *1* | *--* | *--* | *--* | *1* |
| *Seizure* | *2* | *--* | *--* | *--* | *2* |
| **Psychiatric disorders** | **2** | **4** | **--** | **--** | **6** |
| *Agitation* | *1* | *1* | *--* | *--* | *2* |
| *Confusion* | *--* | *1* | *--* | *--* | *1* |
| *Depression* | *1* | *1* | *--* | *--* | *2* |
| *Suicidal ideation* | *--* | *1* | *--* | *--* | *1* |
| **Respiratory, thoracic and mediastinal disorders** | **1** | **--** | **--** | **--** | **1** |
| *Sore throat* | *1* | *--* | *--* | *--* | *1* |
| **Skin and subcutaneous tissue disorders** | **2** | **1** | **--** | **--** | **3** |
| *Alopecia* | *--* | *1* | *--* | *--* | *1* |
| *Pruritus* | *1* | *--* | *--* | *--* | *1* |
| *Rash maculo-papular* | 1 | -- | -- | -- | 1 |
| **Cohort 2** | **28** | **11** | **1** | **--** | **40** |
| **Blood and lymphatic system disorders** | **1** | **--** | **--** | **--** | **1** |
| *Lymph node pain* | 1 | -- | -- | -- | 1 |
| **Ear and labyrinth disorders** | **--** | **1** | **--** | **--** | **1** |
| *Hearing impaired* | -- | 1 | -- | -- | 1 |
| **Eye disorders** | **1** | **--** | **--** | **--** | **1** |
| *Vision decreased* | 1 | -- | -- | -- | 1 |
| **Gastrointestinal disorders** | **1** | **1** | **--** | **--** | **2** |
| *Diarrhea* | 1 | -- | -- | -- | 1 |
| *Nausea* | -- | 1 | -- | -- | 1 |
| **General disorders and administration site conditions** | **6** | **--** | **--** | **--** | **6** |
| *Flu-like symptoms* | 1 | -- | -- | -- | 1 |
| *Gait disturbance* | 1 | -- | -- | -- | 1 |
| *Injection site reaction* | 3 | -- | -- | -- | 3 |
| *Non-cardiac chest pain* | 1 | -- | -- | -- | 1 |
| **Investigations** | **--** | **4** | **--** | **--** | **4** |
| *Lymphocyte count decreased* | -- | 2 | -- | -- | 2 |
| *Neutrophil count decreased* | -- | 1 | -- | -- | 1 |
| *Weight loss* | -- | 1 | -- | -- | 1 |
| **Musculoskeletal and connective tissue disorders** | **8** | **--** | **--** | **--** | **8** |
| *Arthralgia* | 2 | -- | -- | -- | 2 |
| *Back Pain* | 1 | -- | -- | -- | 1 |
| *Generalized muscle weakness* | 1 | -- | -- | -- | 1 |
| *Myalgia* | 2 | -- | -- | -- | 2 |
| *Neck pain* | 2 | -- | -- | -- | 2 |
| **Nervous system disorders** | **3** | **1** | **1** | **--** | **5** |
| *Concentration Impairment* | 1 | -- | -- | -- | 1 |
| *Dizziness* | 2 | -- | -- | -- | 2 |
| *Encephalopathy* | -- | -- | 1 | -- | 1 |
| *Lethargy* | -- | 1 | -- | -- | 1 |
| **Psychiatric disorders** | **2** | **1** | **--** | **--** | **3** |
| *Confusion* | 1 | 1 | -- | -- | 2 |
| *Depression* | 1 | -- | -- | -- | 1 |
| **Renal and urinary disorders** | **1** | **--** | **--** | **--** | **1** |
| *Urinary tract pain* | 1 | -- | -- | -- | 1 |
| **Skin and subcutaneous tissue disorders** | **5** | **3** | **--** | **--** | **8** |
| *Alopecia* | -- | 1 | -- | -- | 1 |
| *Pruritus* | 1 | -- | -- | -- | 1 |
| *Rash maculo-papular* | 3 | -- | -- | -- | 3 |
| *Urticaria* | 1 | 2 | -- | -- | 3 |
| **Cohort 3** | **30** | **6** | **--** | **--** | **36** |
| **Blood and lymphatic system disorders** | **1** | **--** | **--** | **--** | **1** |
| *Eosinophilia* | 1 | -- | -- | -- | 1 |
| **Ear and labyrinth disorders** | **1** | -- | -- | -- | **1** |
| *Tinnitus* | 1 | -- | -- | -- | 1 |
| **Eye disorders** | **1** | -- | -- | -- | **1** |
| *Eye disorders - Other, specify* | 1 | -- | -- | -- | 1 |
| **Gastrointestinal disorders** | **4** | -- | -- | -- | **4** |
| *Constipation* | 1 | -- | -- | -- | 1 |
| *Diarrhea* | 2 | -- | -- | -- | 2 |
| *Nausea* | 1 | -- | -- | -- | 1 |
| **General disorders and administration site conditions** | **7** | **2** | -- | -- | **9** |
| *Fatigue* | 1 | 1 | -- | -- | 2 |
| *Gait disturbance* | 1 | -- | -- | -- | 1 |
| *Injection site reaction* | 4 | 1 | -- | -- | 5 |
| *Localized edema* | 1 | -- | -- | -- | 1 |
| **Infections and infestations** | -- | **1** | -- | -- | **1** |
| *Thrush* | -- | 1 | -- | -- | 1 |
| **Injury, poisoning and procedural complications** | **1** | -- | -- | -- | **1** |
| *Fall* | 1 | -- | -- | -- | 1 |
| **Musculoskeletal and connective tissue disorders** | **5** | **1** | -- | -- | **6** |
| *Back Pain* | -- | 1 | -- | -- | 1 |
| *Myalgia* | 1 | -- | -- | -- | 1 |
| *Neck pain* | 4 | -- | -- | -- | 4 |
| **Nervous system disorders** | **4** | **1** | -- | -- | **5** |
| *Dizziness* | 2 | -- | -- | -- | 2 |
| *Headache* | -- | 1 | -- | -- | 1 |
| *Paresthesia* | 1 | -- | -- | -- | 1 |
| *Seizure* | 1 | -- | -- | -- | 1 |
| **Psychiatric disorders** | **1** | -- | -- | -- | **1** |
| *Confusion* | 1 | -- | -- | -- | 1 |
| **Skin and subcutaneous tissue disorders** | **5** | **1** | -- | -- | **6** |
| *Hyperhidrosis* | 1 | -- | -- | -- | 1 |
| *Pruritus* | 2 | -- | -- | -- | 2 |
| *Urticaria* | 2 | 1 | -- | -- | 3 |
| **Cohort 4** | **39** | **9** | **4** | **1** | **53** |
| **Ear and labyrinth disorders** | **2** | -- | -- | -- | **2** |
| *Tinnitus* | 2 | -- | -- | -- | 2 |
| **Eye disorders** | **2** | -- | -- | -- | **2** |
| *Eye disorders - Other, specify* | 1 | -- | -- | -- | 1 |
| *Periorbital edema* | 1 | -- | -- | -- | 1 |
| **Gastrointestinal disorders** | **5** | **1** | -- | -- | **6** |
| *Abdominal pain* | 2 | -- | -- | -- | 2 |
| *Constipation* | 1 | -- | -- | -- | 1 |
| *Diarrhea* | *1* | -- | -- | -- | *1* |
| *Nausea* | *1* | *1* | -- | -- | *2* |
| **General disorders and administration site conditions** | **6** | -- | -- | -- | **6** |
| *Fatigue* | *3* | -- | -- | -- | *3* |
| *Flu-like symptoms* | *1* | -- | -- | -- | *1* |
| *Injection site reaction* | *2* | -- | -- | -- | *2* |
| **Immune system disorders** | **1** | -- | -- | -- | **1** |
| *Autoimmune disorder* | *1* | -- | -- | -- | *1* |
| **Infections and infestations** | **1** | -- | -- | -- | **1** |
| *Thrush* | *1* | -- | -- | -- | *1* |
| **Injury, poisoning and procedural complications** | **1** | -- | -- | -- | **1** |
| *Bruising* | *1* | -- | -- | -- | *1* |
| **Investigations** | **2** | **3** | **3** | **1** | **9** |
| *Neutrophil count decreased* | -- | *1* | *1* | *1* | *3* |
| *Platelet count decreased* | *2* | *1* | *1* | -- | *4* |
| *White blood cell decreased* | -- | *1* | *1* | -- | *2* |
| **Metabolism and nutrition disorders** | **1** | -- | -- | -- | **1** |
| *Anorexia* | *1* | -- | -- | -- | *1* |
| **Musculoskeletal and connective tissue disorders** | **3** | **1** | -- | -- | **4** |
| *Myalgia* | -- | *1* | -- | -- | *1* |
| *Neck pain* | *3* | -- | -- | -- | *3* |
| **Nervous system disorders** | **6** | **4** | **1** | -- | **11** |
| *Dizziness* | *1* | -- | -- | -- | *1* |
| *Dysgeusia* | *2* | *1* | -- | -- | *3* |
| *Headache* | *2* | *2* | -- | -- | *4* |
| *Paresthesia* | *1* | *1* | -- | -- | *2* |
| *Seizure* | -- | -- | *1* | -- | *1* |
| **Reproductive system and breast disorders** | **1** | -- | -- | -- | **1** |
| *Irregular menstruation* | *1* | -- | -- | -- | *1* |
| **Respiratory, thoracic and mediastinal disorders** | **1** | -- | -- | -- | **1** |
| *Post Nasal Drip* | *1* | -- | -- | -- | *1* |
| **Skin and subcutaneous tissue disorders** | **6** | -- | -- | -- | **6** |
| *Pruritus* | *4* | -- | -- | -- | *4* |
| *Skin hyperpigmentation* | *1* | -- | -- | -- | *1* |
| *Urticaria* | *1* | -- | -- | -- | *1* |
| **Skin and subcutaneous tissue disorders - Other, specify** | **1** | -- | -- | -- | **1** |
| *Skin and subcutaneous tissue disorders - Other, specify* | *1* | -- | -- | -- | *1* |
| **Grand Total** | **137** | **49** | **7** | **1** | **194** |

**Table S2.** Operative Covariates.

| **Variable** | **Reoperation** | **No Reoperation** |
| --- | --- | --- |
| Age | 58 | 63 |
| Female Sex | 14% (1/7) | 78% (7/9) |
| Baseline ECOG=0 | 43% | 33% |
| GTR/NTR | 86% (6/7) | 67% (6/9) |
| MGMT Unmethylated | 100.0% (7/7) | 88.9% (8/9) |

