## Supplemental Figures File for "Phase I Clinical Study of DOC1021 (dubodencel) for Adjuvant Immunotherapy of Glioblastoma"

Figure S1

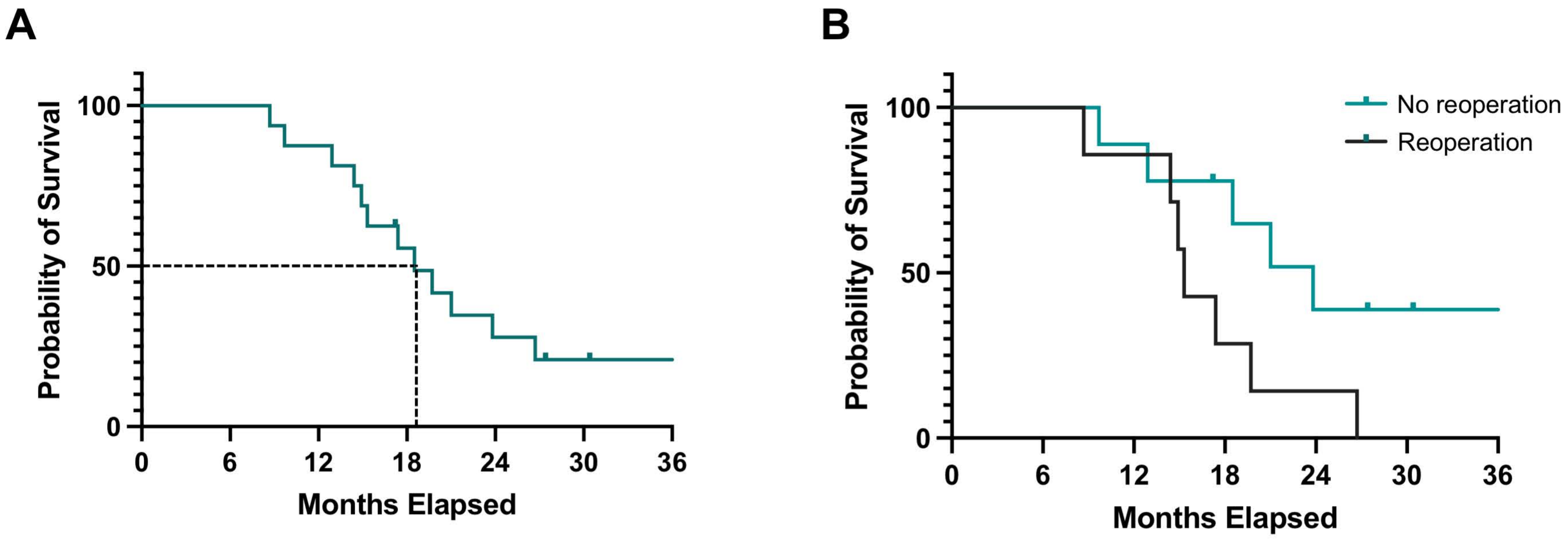
A
B
Probability of Survival
100
50
0
0
6
12
18
24
30
36
Months Elapsed
No reoperation
Reoperation

Figure S2

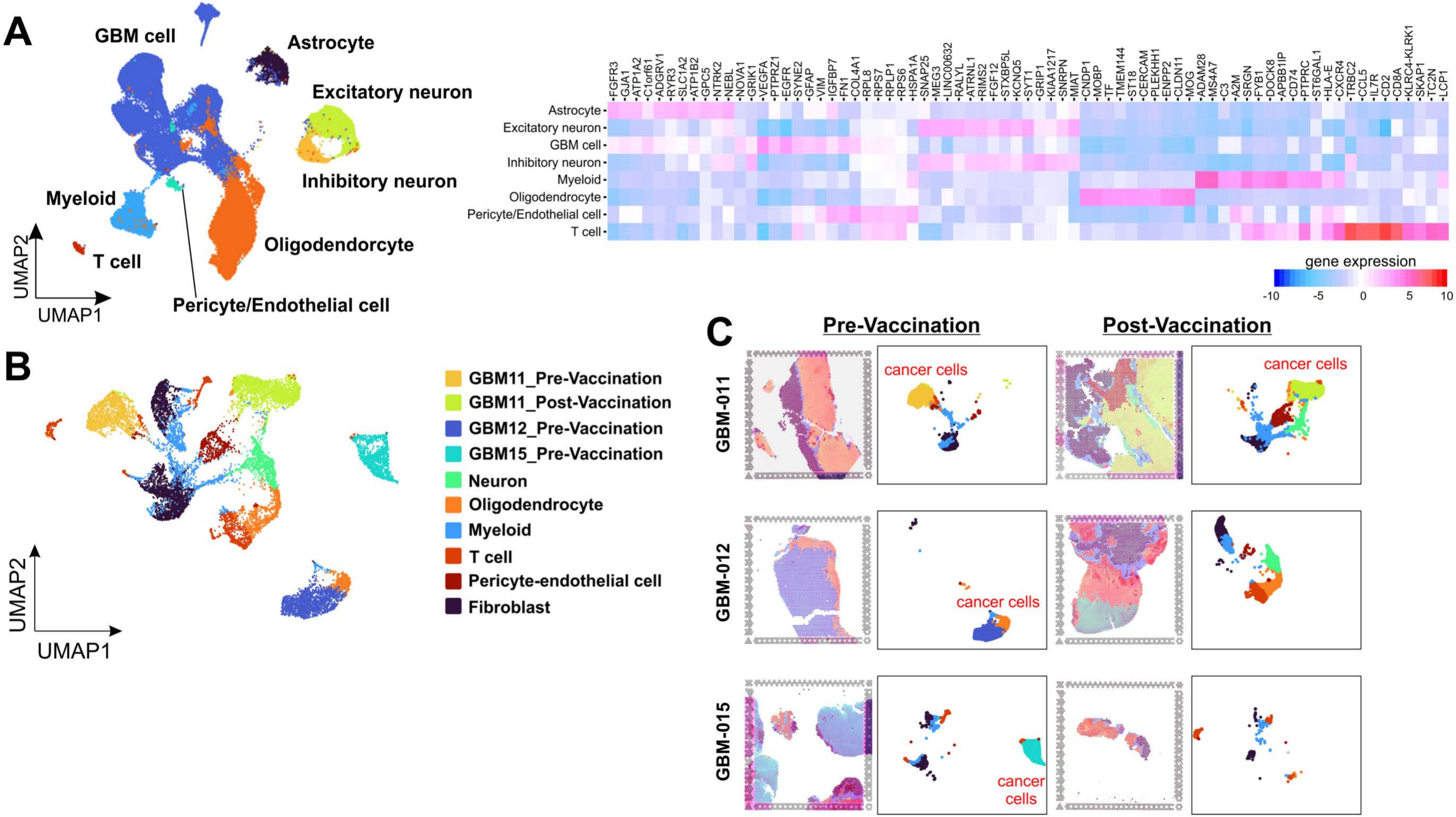
A
GBM cell
Astrocyte
Excitatory neuron
Inhibitory neuron
Myeloid
Oligodendorcyte
T cell
Pericyte/Endothelial cell
UMAP1
UMAP2
Astrocyte
Excitatory neuron
GBM cell
Inhibitory neuron
Myeloid
Oligodendrocyte
Pericyte/Endothelial cell
T cell
gene expression
-10 -5 0 5 10
B
GBM11_Pre-Vaccination
GBM11_Post-Vaccination
GBM12_Pre-Vaccination
GBM15_Pre-Vaccination
Neuron
Oligodendrocyte
Myeloid
T cell
Pericyte-endothelial cell
Fibroblast
UMAP1
UMAP2
C
Pre-Vaccination
Post-Vaccination
GBM-011
GBM-012
GBM-015
cancer cells

Figure S3

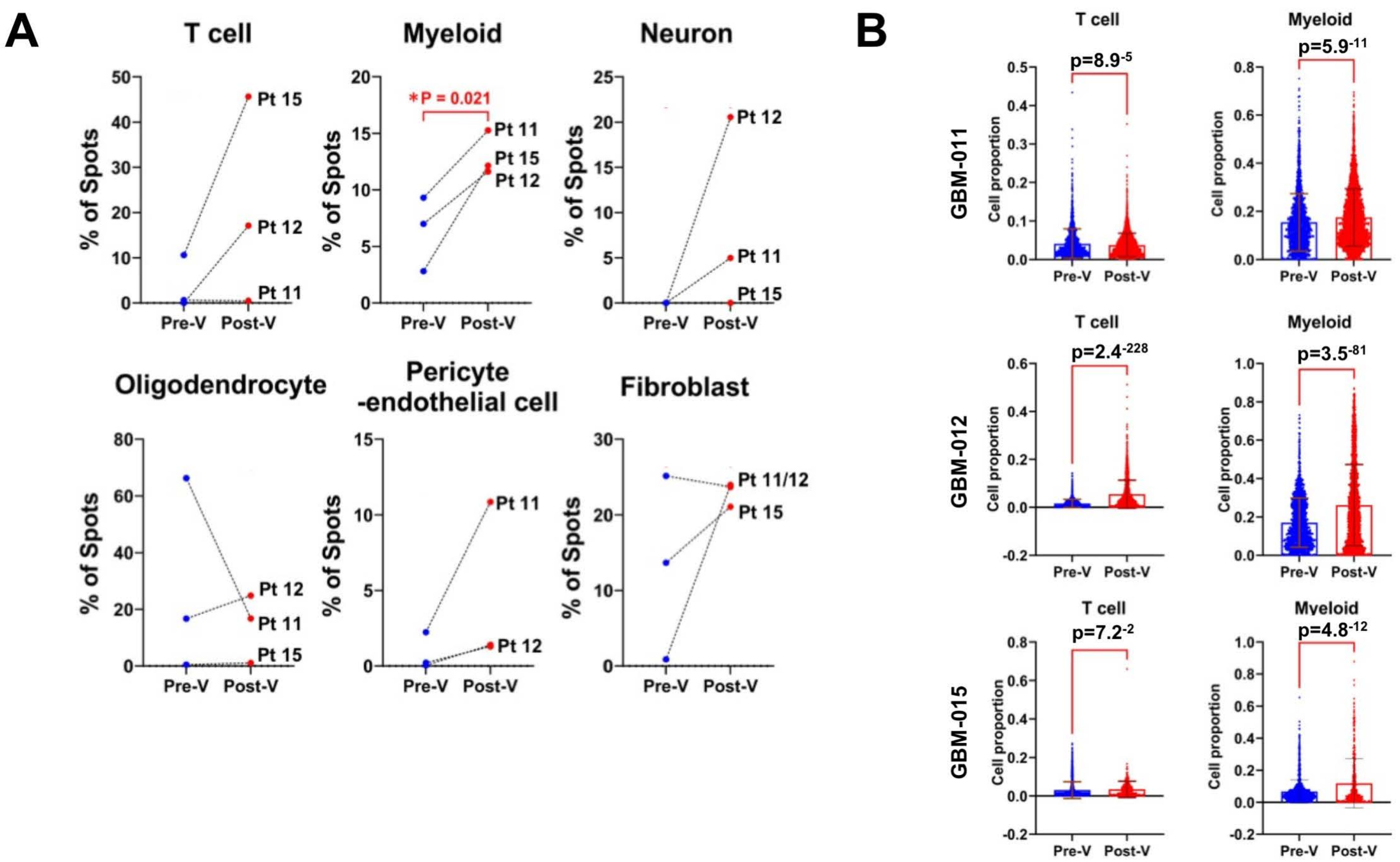

A
T cell
Myeloid
Neuron
% of Spots
Pre-V
Post-V
Pt 15
Pt 12
Pt 11
*P = 0.021
Oligodendrocyte
Pericyte
-endothelial cell
Fibroblast
Pt 11/12
B
GBM-011
GBM-012
GBM-015
Cell proportion
p=8.9-5
p=5.9-11
p=2.4-228
p=3.5-81
p=7.2-2
p=4.8-12

Figure S4

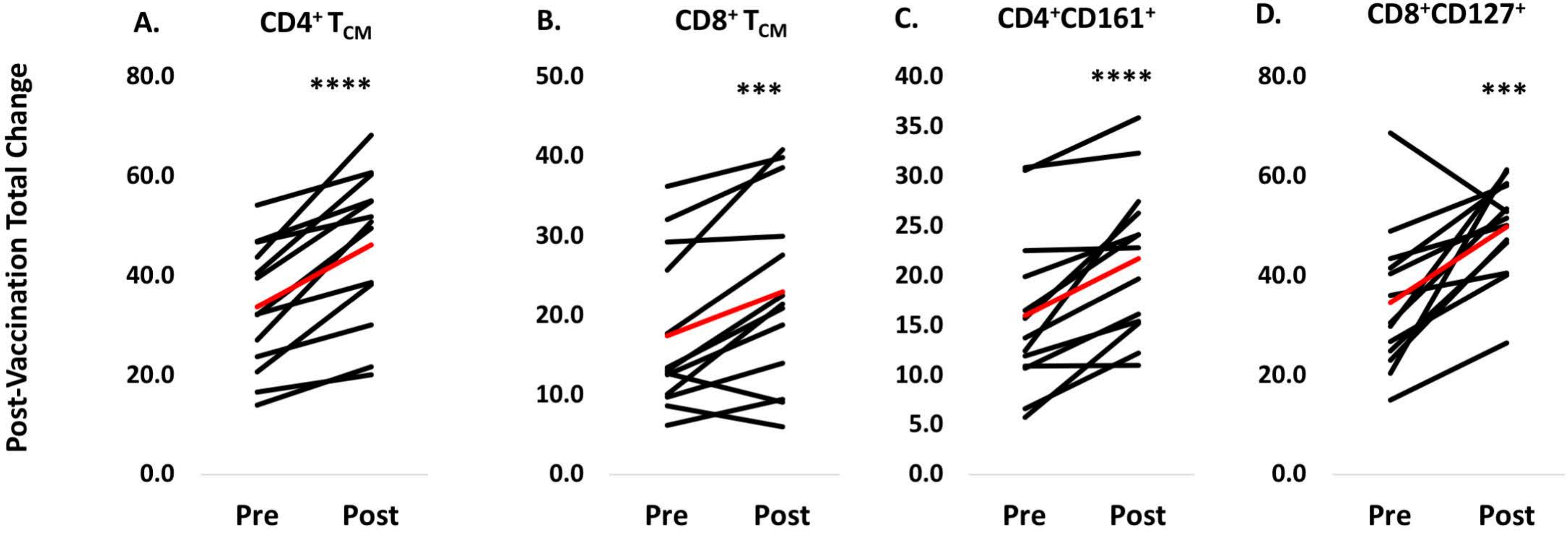
A.
CD4+ TCM
B.
CD8+ TCM
C.
CD4+CD161+
D.
CD8+CD127+
Post-Vaccination Total Change
80.0
60.0
40.0
20.0
0.0
****
50.0
40.0
30.0
20.0
10.0
0.0
***
40.0
35.0
30.0
25.0
20.0
15.0
10.0
5.0
0.0
****
80.0
60.0
40.0
20.0
0.0
***
Pre
Post
Pre
Post
Pre
Post
Pre
Post

Figure S5

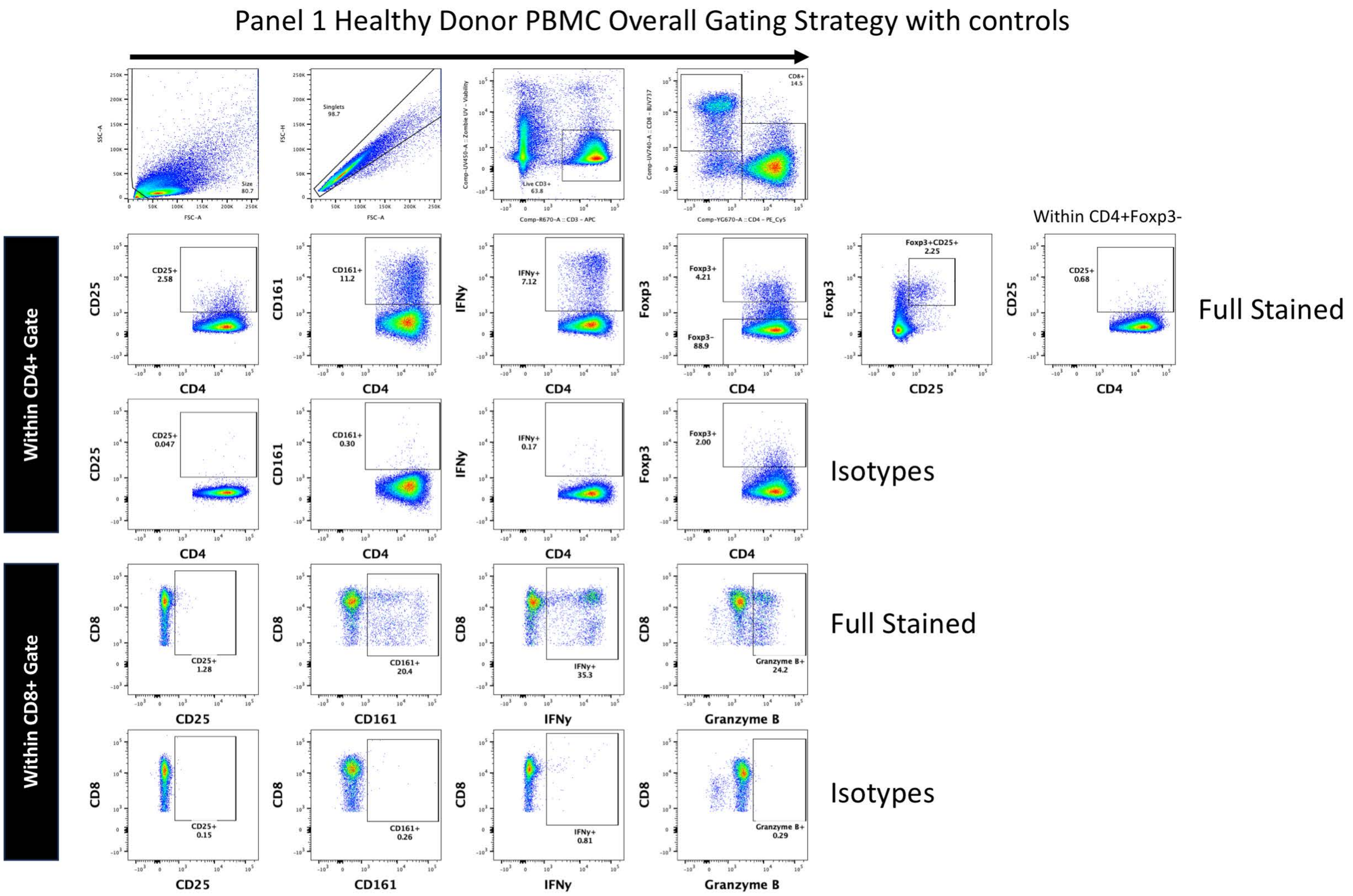

Panel 1 Healthy Donor PBMC Overall Gating Strategy with controls
Size 80.7
Singlets 98.7
Live CD3+ 63.8
CD8+ 14.5
Within CD4+ Gate
Within CD8+ Gate
Within CD4+Foxp3-
CD25+ 2.58
CD161+ 11.2
IFNγ+ 7.12
Foxp3+ 4.21
Foxp3- 88.9
Foxp3+CD25+ 2.25
CD25+ 0.68
Full Stained
CD25+ 0.047
CD161+ 0.30
IFNγ+ 0.17
Foxp3+ 2.00
Isotypes
CD25+ 1.28
CD161+ 20.4
IFNγ+ 35.3
Granzyme B+ 24.2
Full Stained
CD25+ 0.15
CD161+ 0.26
IFNγ+ 0.81
Granzyme B+ 0.29
Isotypes
CD4
CD25
CD161
IFNγ
Foxp3
CD8
Granzyme B

Figure S6

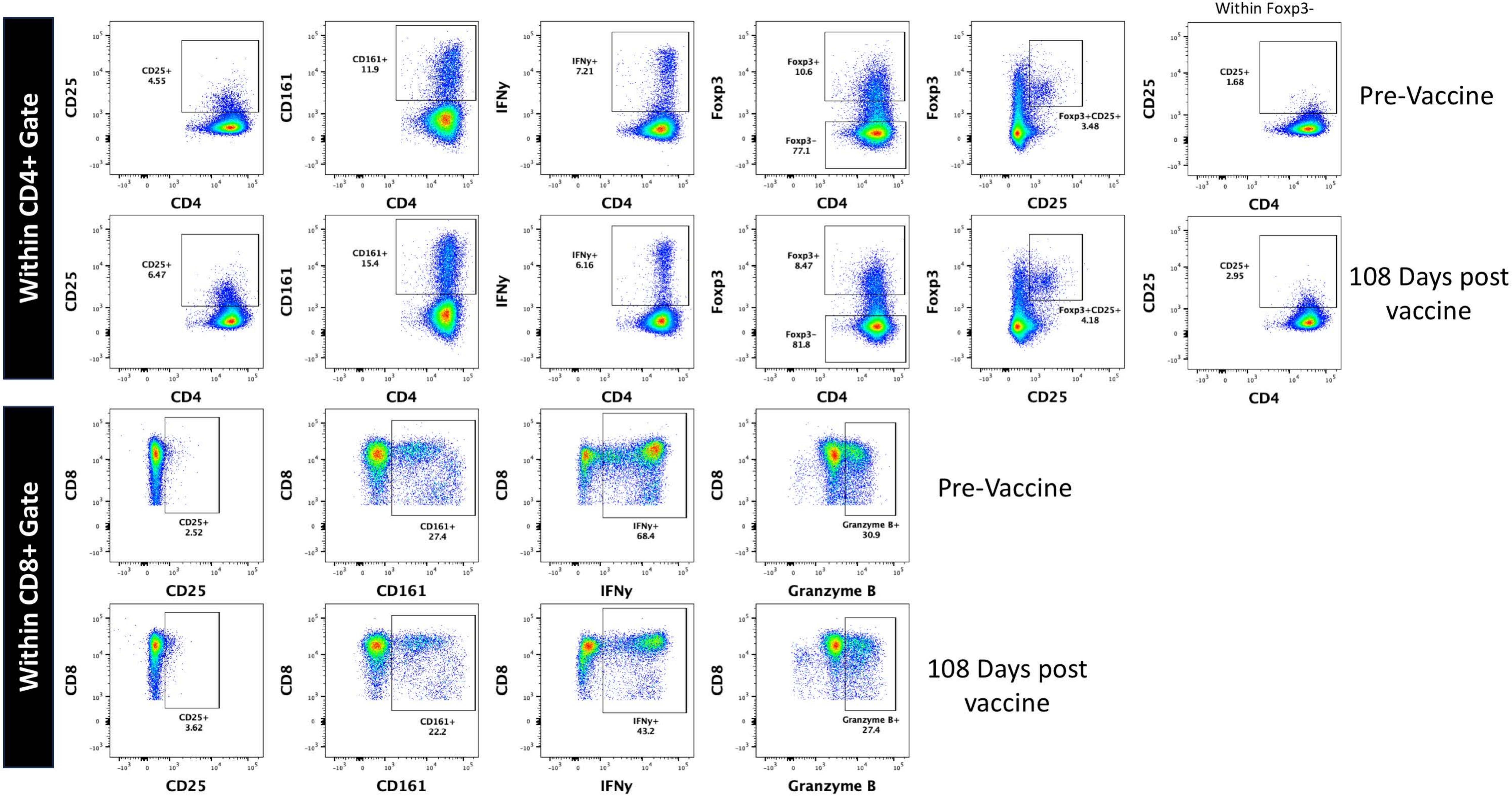

Panel 1 GBM-011
Within CD4+ Gate
Within CD8+ Gate
Within Foxp3-
Pre-Vaccine
108 Days post vaccine
CD25+ 4.55
CD161+ 11.9
IFNy+ 7.21
Foxp3+ 10.6
Foxp3- 77.1
Foxp3+CD25+ 3.48
CD25+ 1.68
CD25+ 6.47
CD161+ 15.4
IFNy+ 6.16
Foxp3+ 8.47
Foxp3- 81.8
Foxp3+CD25+ 4.18
CD25+ 2.95
CD25+ 2.52
CD161+ 27.4
IFNy+ 68.4
Granzyme B+ 30.9
CD25+ 3.62
CD161+ 22.2
IFNy+ 43.2
Granzyme B+ 27.4
CD4
CD25
CD161
IFNy
Foxp3
CD8
Granzyme B

Figure S7

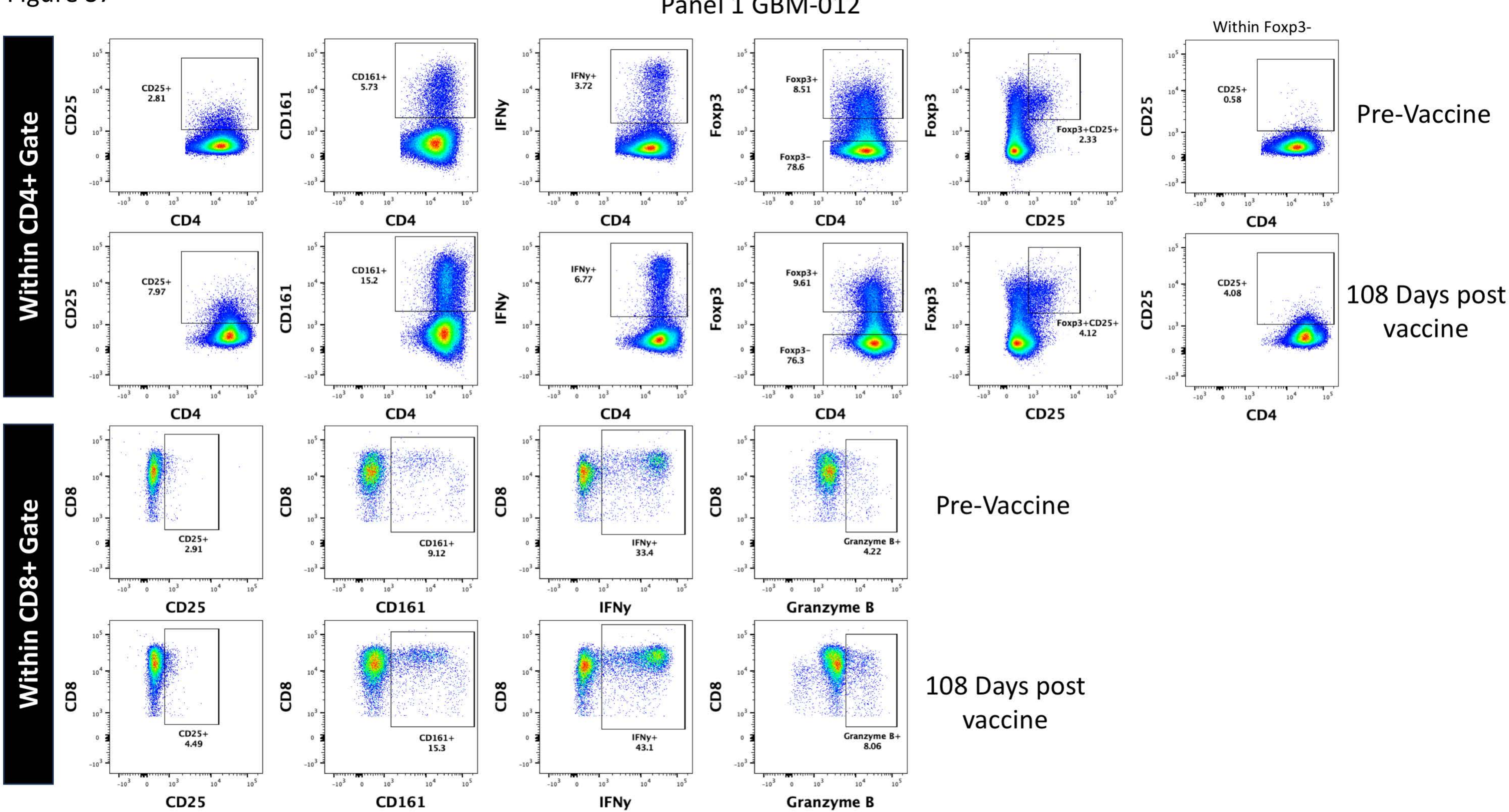

Panel 1 GBM-012
Within CD4+ Gate
Within CD8+ Gate
Within Foxp3-
Pre-Vaccine
108 Days post vaccine
CD25+ 2.81
CD161+ 5.73
IFNy+ 3.72
Foxp3+ 8.51
Foxp3- 78.6
Foxp3+CD25+ 2.33
CD25+ 0.58
CD25+ 7.97
CD161+ 15.2
IFNy+ 6.77
Foxp3+ 9.61
Foxp3- 76.3
Foxp3+CD25+ 4.12
CD25+ 4.08
CD25+ 2.91
CD161+ 9.12
IFNy+ 33.4
Granzyme B+ 4.22
CD25+ 4.49
CD161+ 15.3
IFNy+ 43.1
Granzyme B+ 8.06
CD4
CD25
CD161
IFNy
Foxp3
CD8
Granzyme B

Figure S8

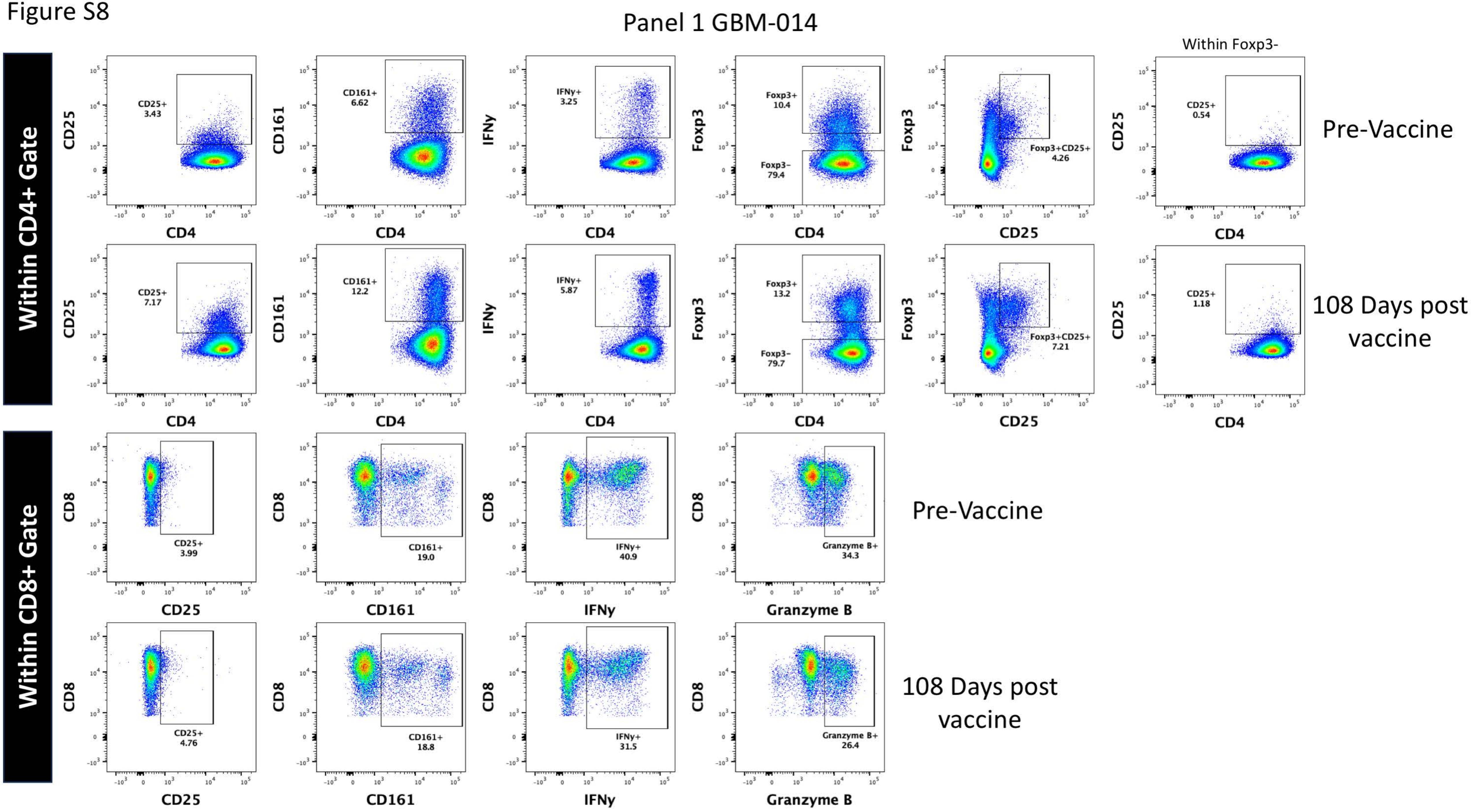

Panel 1 GBM-014
Within CD4+ Gate
Within CD8+ Gate
Within Foxp3-
Pre-Vaccine
108 Days post vaccine
CD25+ 3.43
CD161+ 6.62
IFNy+ 3.25
Foxp3+ 10.4
Foxp3- 79.4
Foxp3+CD25+ 4.26
CD25+ 0.54
CD25+ 7.17
CD161+ 12.2
IFNy+ 5.87
Foxp3+ 13.2
Foxp3- 79.7
Foxp3+CD25+ 7.21
CD25+ 1.18
CD25+ 3.99
CD161+ 19.0
IFNy+ 40.9
Granzyme B+ 34.3
CD25+ 4.76
CD161+ 18.8
IFNy+ 31.5
Granzyme B+ 26.4
CD4
CD25
CD161
IFNy
Foxp3
CD8
Granzyme B

Figure S9

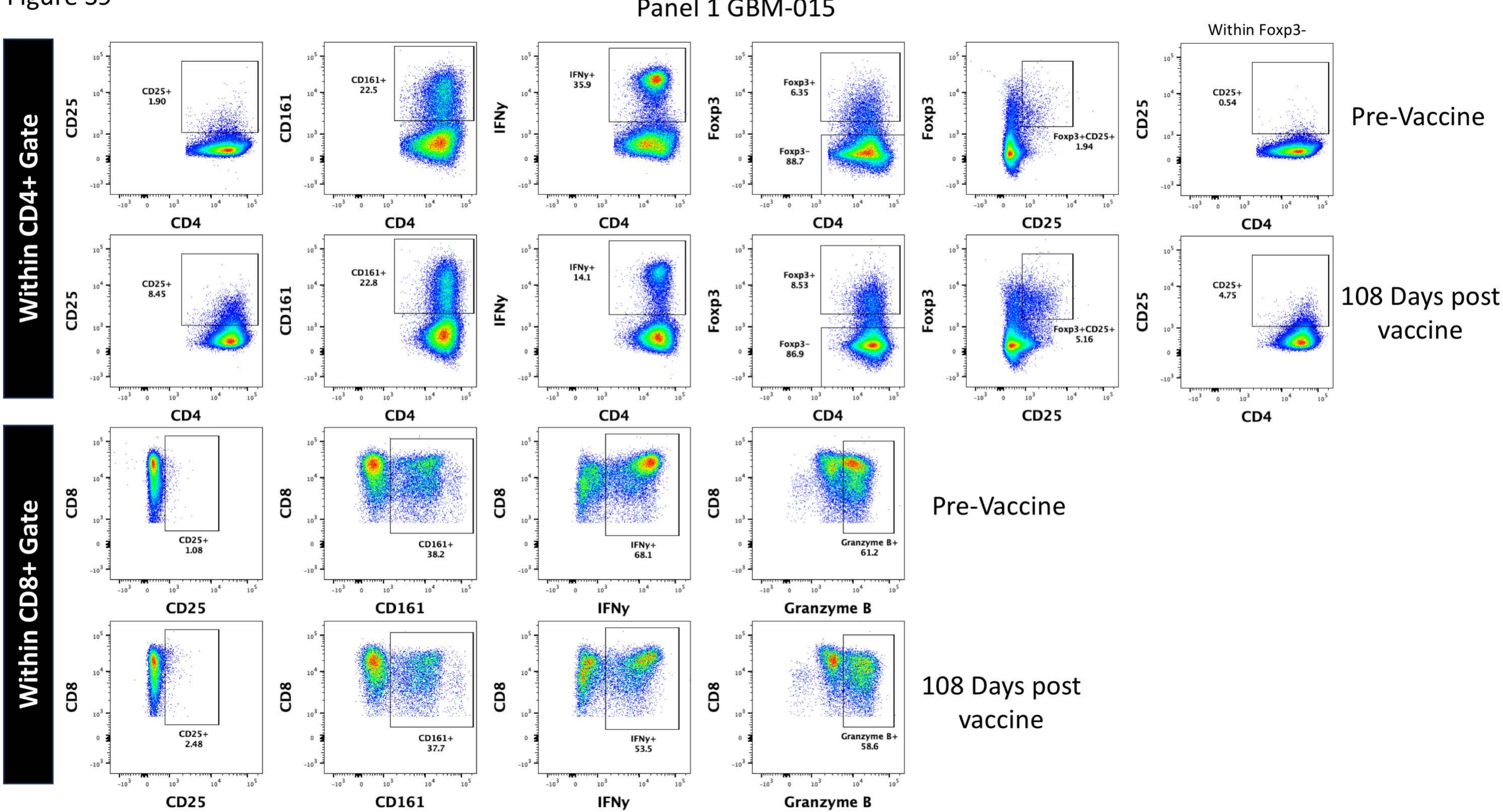
Panel 1 GBM-015
Within CD4+ Gate
Within CD8+ Gate
Within Foxp3-
Pre-Vaccine
108 Days post vaccine
CD25+ 1.90
CD161+ 22.5
IFNγ+ 35.9
Foxp3+ 6.35
Foxp3− 88.7
Foxp3+CD25+ 1.94
CD25+ 0.54
CD25+ 8.45
CD161+ 22.8
IFNγ+ 14.1
Foxp3+ 8.53
Foxp3− 86.9
Foxp3+CD25+ 5.16
CD25+ 4.75
CD25+ 1.08
CD161+ 38.2
IFNγ+ 68.1
Granzyme B+ 61.2
CD25+ 2.48
CD161+ 37.7
IFNγ+ 53.5
Granzyme B+ 58.6
CD4
CD8
CD25
CD161
IFNγ
Foxp3
Granzyme B

Figure S10

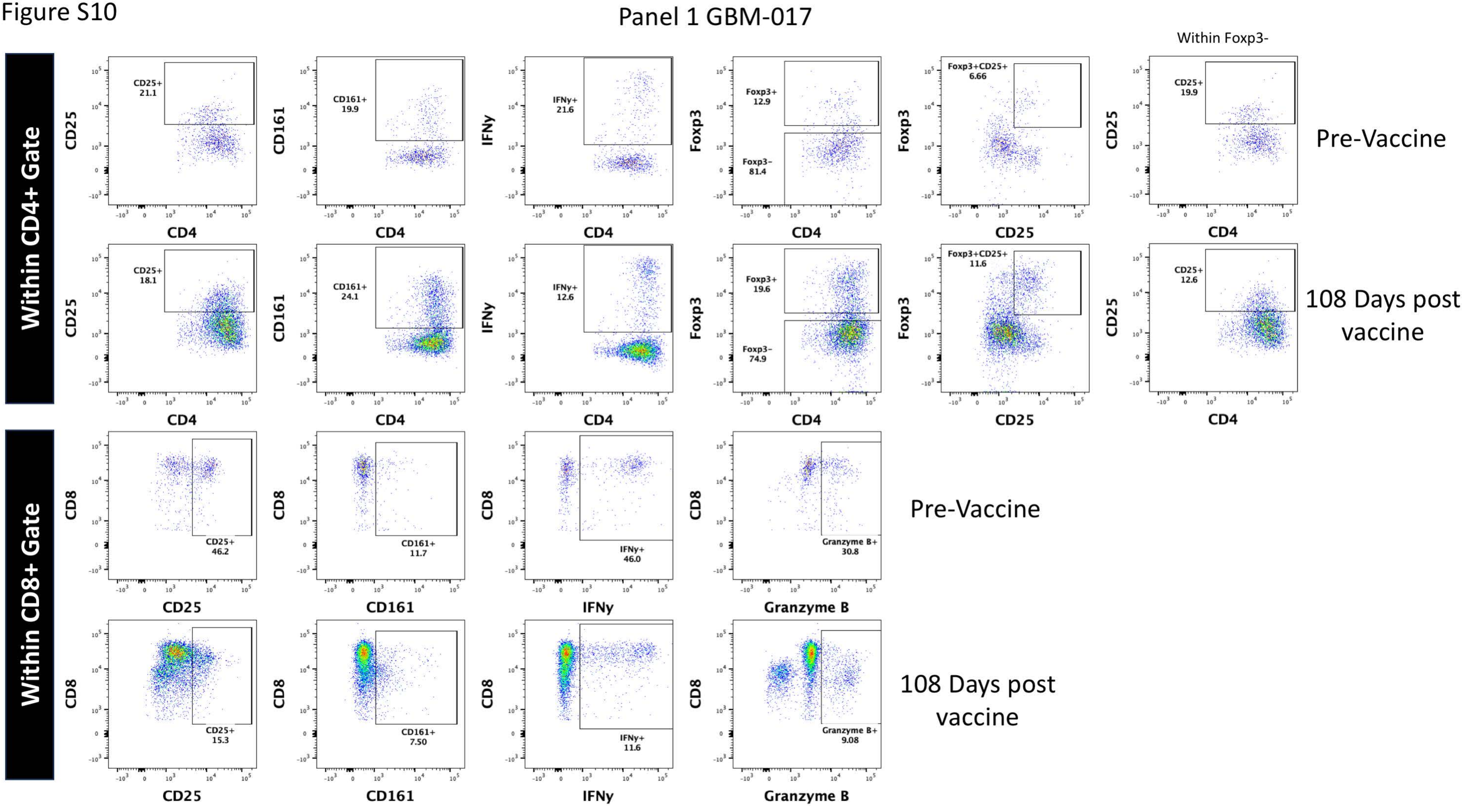
Panel 1 GBM-017
Within CD4+ Gate
Within CD8+ Gate
Within Foxp3-
Pre-Vaccine
108 Days post vaccine
CD25+ 21.1
CD161+ 19.9
IFNy+ 21.6
Foxp3+ 12.9
Foxp3- 81.4
Foxp3+CD25+ 6.66
CD25+ 19.9
CD25+ 18.1
CD161+ 24.1
IFNy+ 12.6
Foxp3+ 19.6
Foxp3- 74.9
Foxp3+CD25+ 11.6
CD25+ 12.6
CD25+ 46.2
CD161+ 11.7
IFNy+ 46.0
Granzyme B+ 30.8
CD25+ 15.3
CD161+ 7.50
IFNy+ 11.6
Granzyme B+ 9.08
CD4
CD25
CD161
IFNy
Foxp3
CD8
Granzyme B

Figure S11

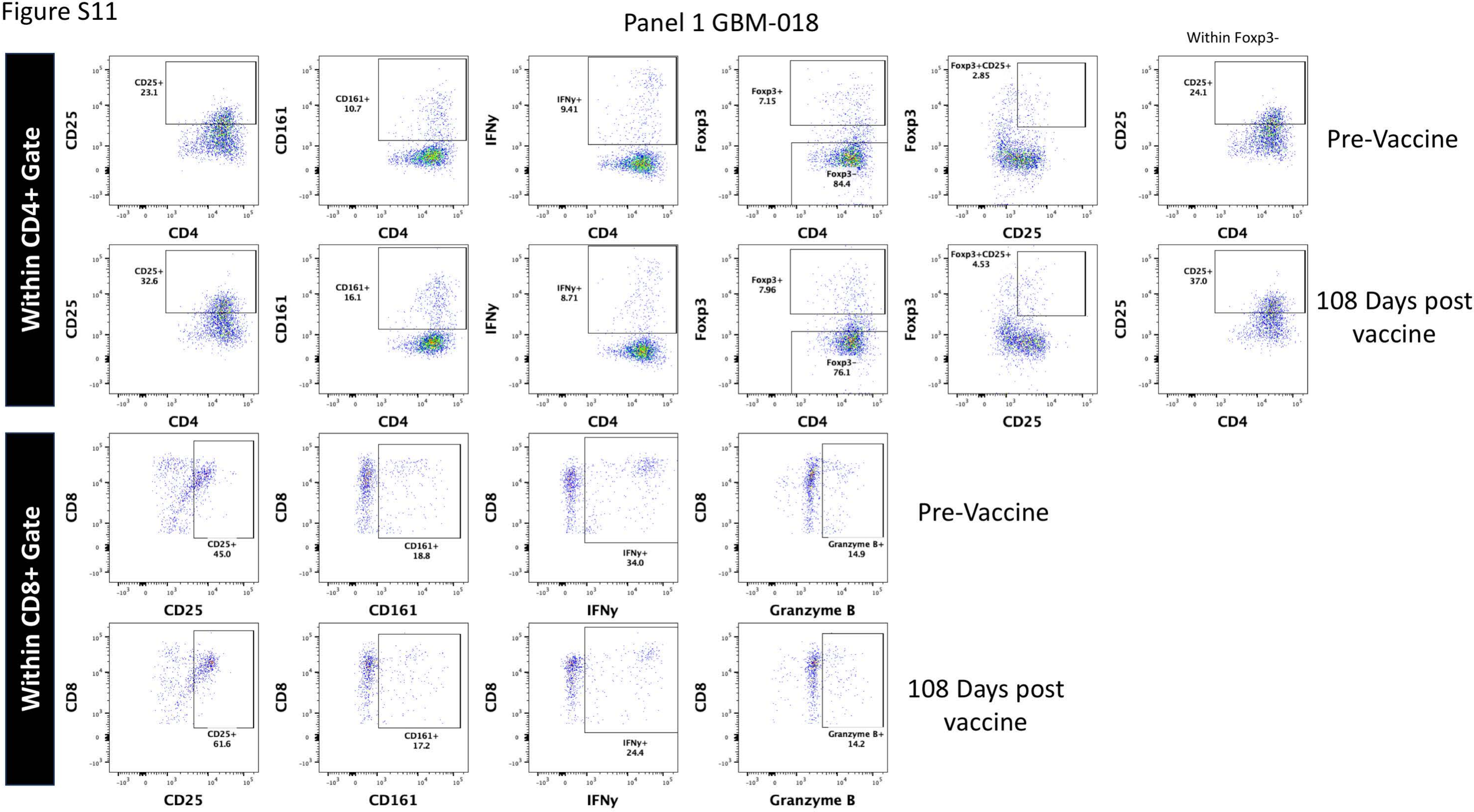
Panel 1 GBM-018
Within Foxp3-
Within CD4+ Gate
Within CD8+ Gate
Pre-Vaccine
108 Days post vaccine
CD25+ 23.1
CD161+ 10.7
IFNy+ 9.41
Foxp3+ 7.15
Foxp3- 84.4
Foxp3+CD25+ 2.85
CD25+ 24.1
CD25+ 32.6
CD161+ 16.1
IFNy+ 8.71
Foxp3+ 7.96
Foxp3- 76.1
Foxp3+CD25+ 4.53
CD25+ 37.0
CD25+ 45.0
CD161+ 18.8
IFNy+ 34.0
Granzyme B+ 14.9
CD25+ 61.6
CD161+ 17.2
IFNy+ 24.4
Granzyme B+ 14.2
CD4
CD25
CD161
IFNy
Foxp3
CD8
Granzyme B

Figure S12

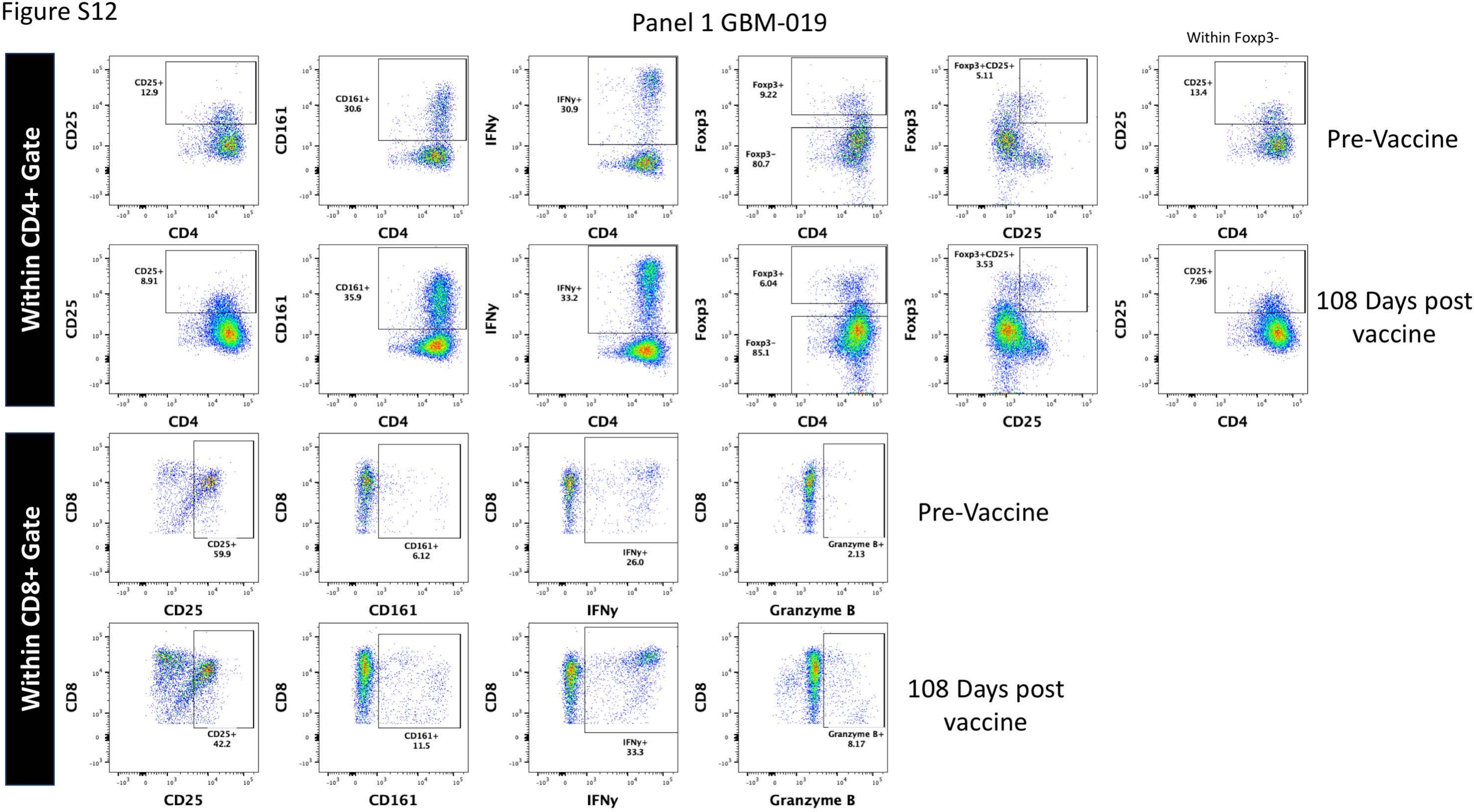

Panel 1 GBM-019
Within Foxp3-
Within CD4+ Gate
Pre-Vaccine
CD25+ 12.9
CD161+ 30.6
IFNy+ 30.9
Foxp3+ 9.22
Foxp3- 80.7
Foxp3+CD25+ 5.11
CD25+ 13.4
108 Days post vaccine
CD25+ 8.91
CD161+ 35.9
IFNy+ 33.2
Foxp3+ 6.04
Foxp3- 85.1
Foxp3+CD25+ 3.53
CD25+ 7.96
CD25
CD161
IFNy
Foxp3
CD4
Within CD8+ Gate
Pre-Vaccine
CD25+ 59.9
CD161+ 6.12
IFNy+ 26.0
Granzyme B+ 2.13
108 Days post vaccine
CD25+ 42.2
CD161+ 11.5
IFNy+ 33.3
Granzyme B+ 8.17
CD8
Granzyme B

Figure S13

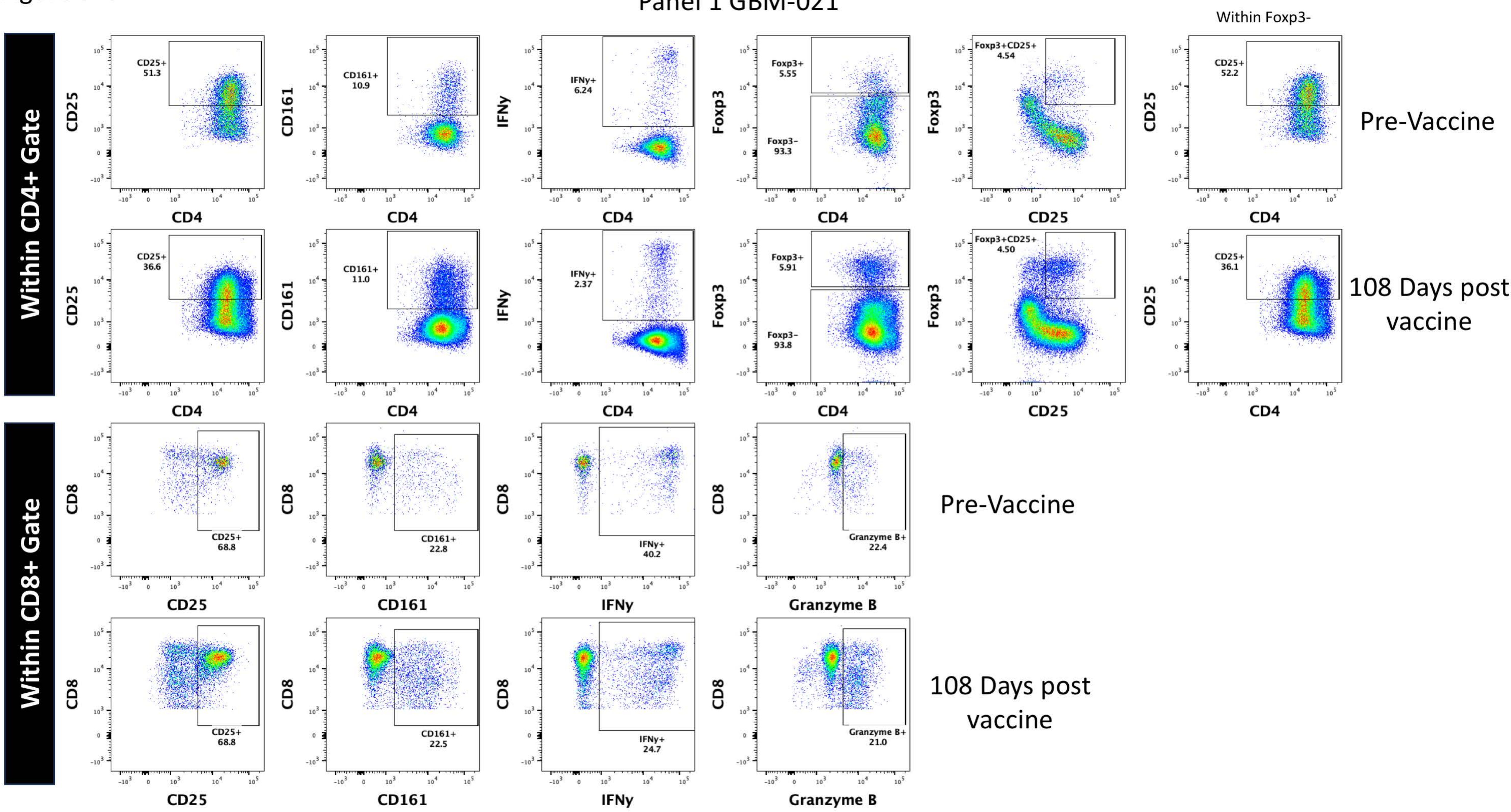

Panel 1 GBM-021
Within Foxp3-
Within CD4+ Gate
Within CD8+ Gate
CD25+ 51.3
CD161+ 10.9
IFNy+ 6.24
Foxp3+ 5.55
Foxp3- 93.3
Foxp3+CD25+ 4.54
CD25+ 52.2
Pre-Vaccine
CD25+ 36.6
CD161+ 11.0
IFNy+ 2.37
Foxp3+ 5.91
Foxp3- 93.8
Foxp3+CD25+ 4.50
CD25+ 36.1
108 Days post vaccine
CD25+ 68.8
CD161+ 22.8
IFNy+ 40.2
Granzyme B+ 22.4
Pre-Vaccine
CD25+ 68.8
CD161+ 22.5
IFNy+ 24.7
Granzyme B+ 21.0
108 Days post vaccine
CD25
CD161
IFNy
Foxp3
CD4
CD8
Granzyme B

Figure S14

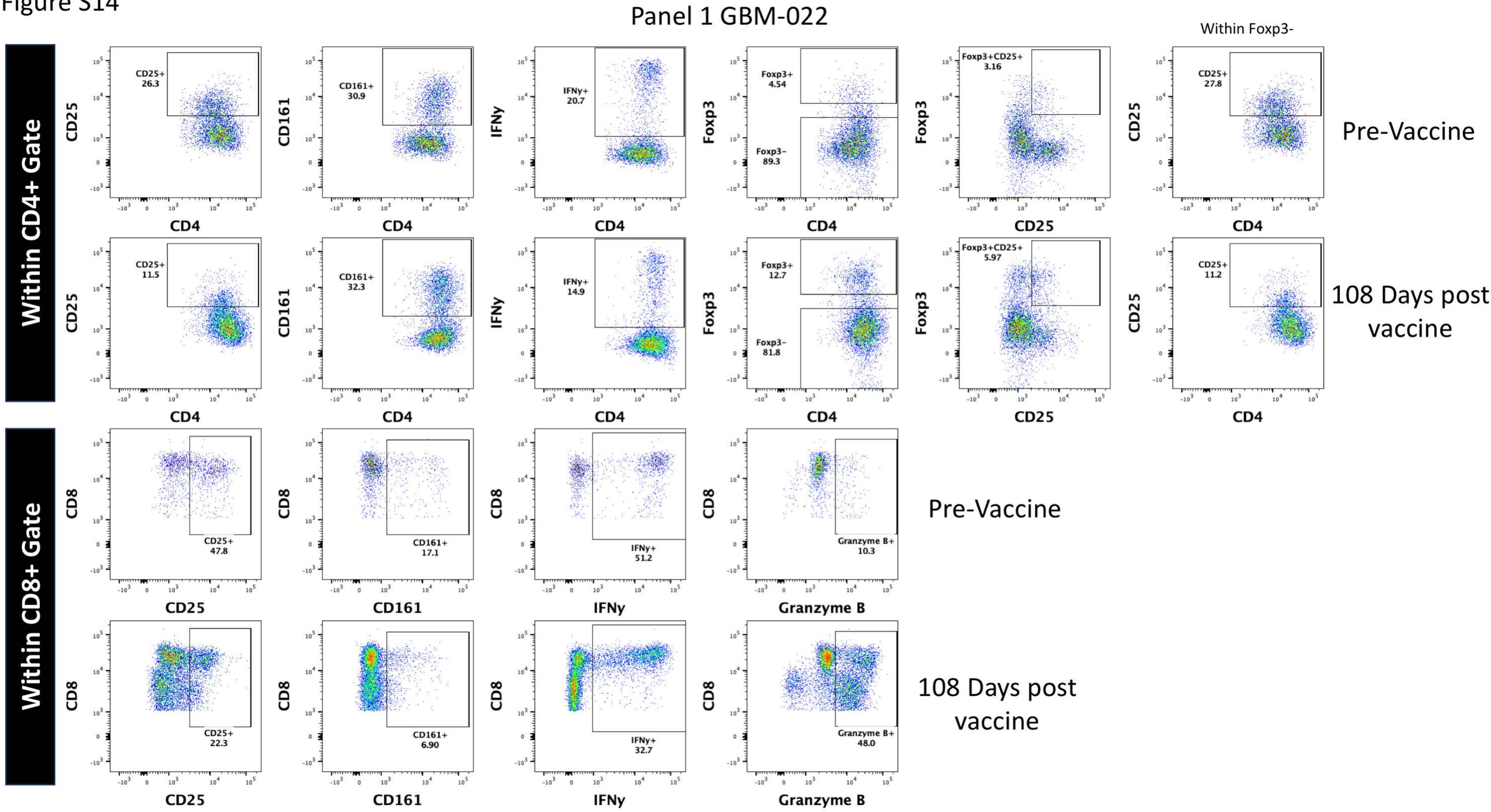
Panel 1 GBM-022
Within Foxp3-
Within CD4+ Gate
Within CD8+ Gate
Pre-Vaccine
108 Days post vaccine
CD25+ 26.3
CD161+ 30.9
IFNy+ 20.7
Foxp3+ 4.54
Foxp3- 89.3
Foxp3+CD25+ 3.16
CD25+ 27.8
CD25+ 11.5
CD161+ 32.3
IFNy+ 14.9
Foxp3+ 12.7
Foxp3- 81.8
Foxp3+CD25+ 5.97
CD25+ 11.2
CD25+ 47.8
CD161+ 17.1
IFNy+ 51.2
Granzyme B+ 10.3
CD25+ 22.3
CD161+ 6.90
IFNy+ 32.7
Granzyme B+ 48.0
CD4
CD25
CD161
IFNy
Foxp3
CD8
Granzyme B

Figure S15

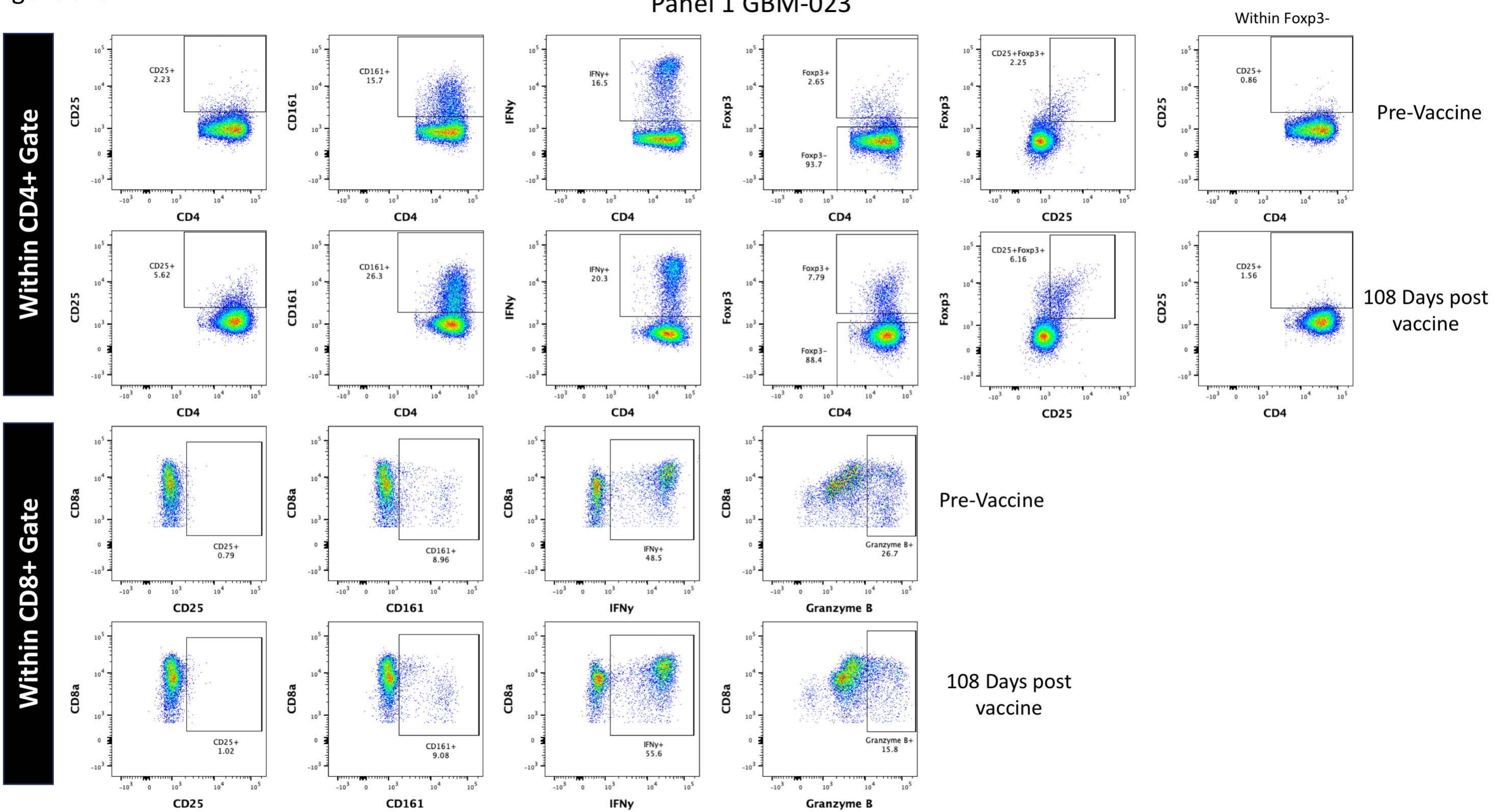

Panel 1 GBM-023
Within CD4+ Gate
Within CD8+ Gate
Within Foxp3-
Pre-Vaccine
108 Days post vaccine
CD25+ 2.23
CD161+ 15.7
IFNγ+ 16.5
Foxp3+ 2.65
Foxp3- 93.7
CD25+Foxp3+ 2.25
CD25+ 0.86
CD25+ 5.62
CD161+ 26.3
IFNγ+ 20.3
Foxp3+ 7.79
Foxp3- 88.4
CD25+Foxp3+ 6.16
CD25+ 1.56
CD25+ 0.79
CD161+ 8.96
IFNγ+ 48.5
Granzyme B+ 26.7
CD25+ 1.02
CD161+ 9.08
IFNγ+ 55.6
Granzyme B+ 15.8
CD4
CD25
CD161
IFNγ
Foxp3
CD8a
Granzyme B

Figure S16

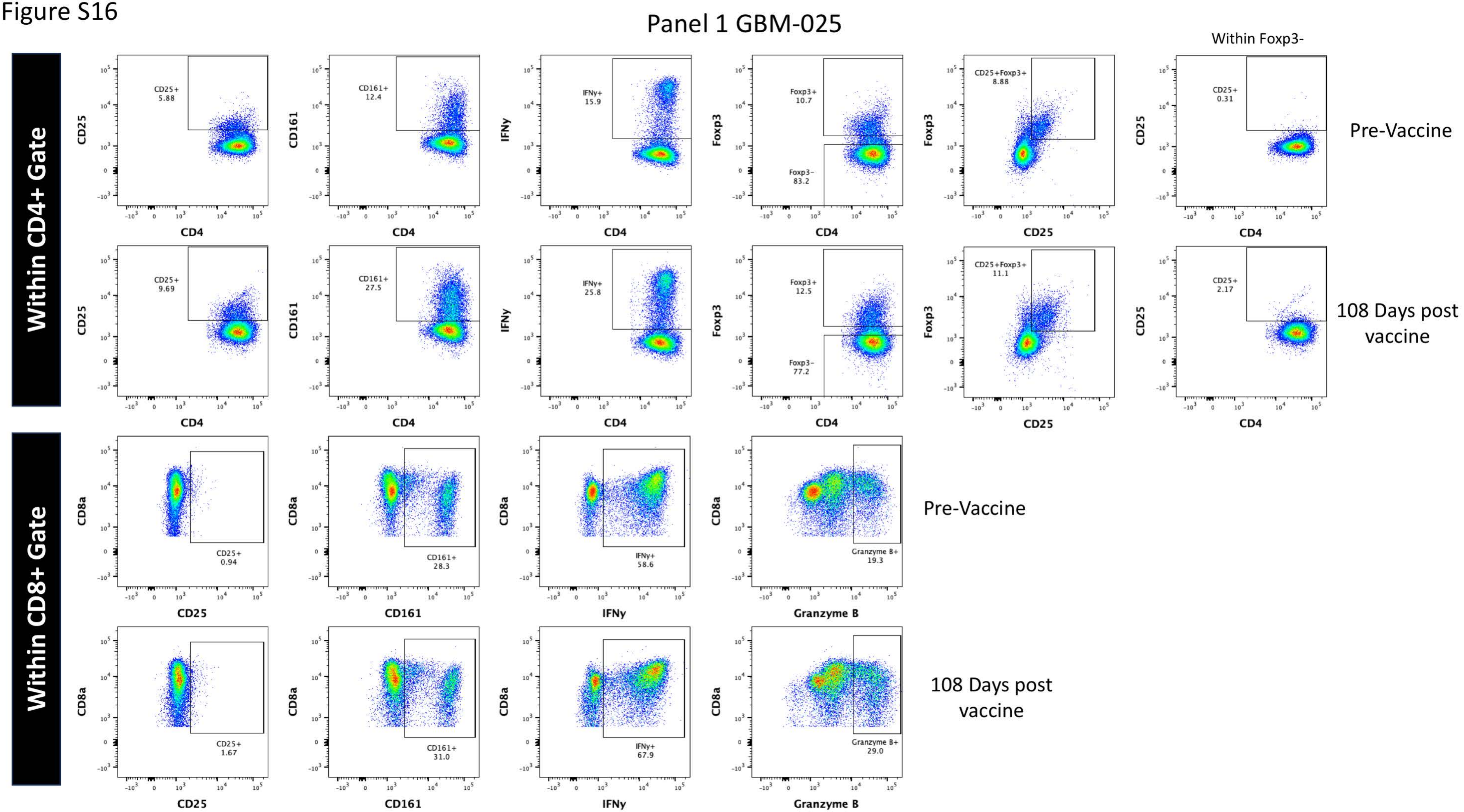
Panel 1 GBM-025
Within CD4+ Gate
Within CD8+ Gate
Within Foxp3-
Pre-Vaccine
108 Days post vaccine
CD25+ 5.88
CD161+ 12.4
IFNγ+ 15.9
Foxp3+ 10.7
Foxp3- 83.2
CD25+Foxp3+ 8.88
CD25+ 0.31
CD25+ 9.69
CD161+ 27.5
IFNγ+ 25.8
Foxp3+ 12.5
Foxp3- 77.2
CD25+Foxp3+ 11.1
CD25+ 2.17
CD25+ 0.94
CD161+ 28.3
IFNγ+ 58.6
Granzyme B+ 19.3
CD25+ 1.67
CD161+ 31.0
IFNγ+ 67.9
Granzyme B+ 29.0
CD4
CD25
CD161
IFNγ
Foxp3
CD8a
Granzyme B

Figure S17

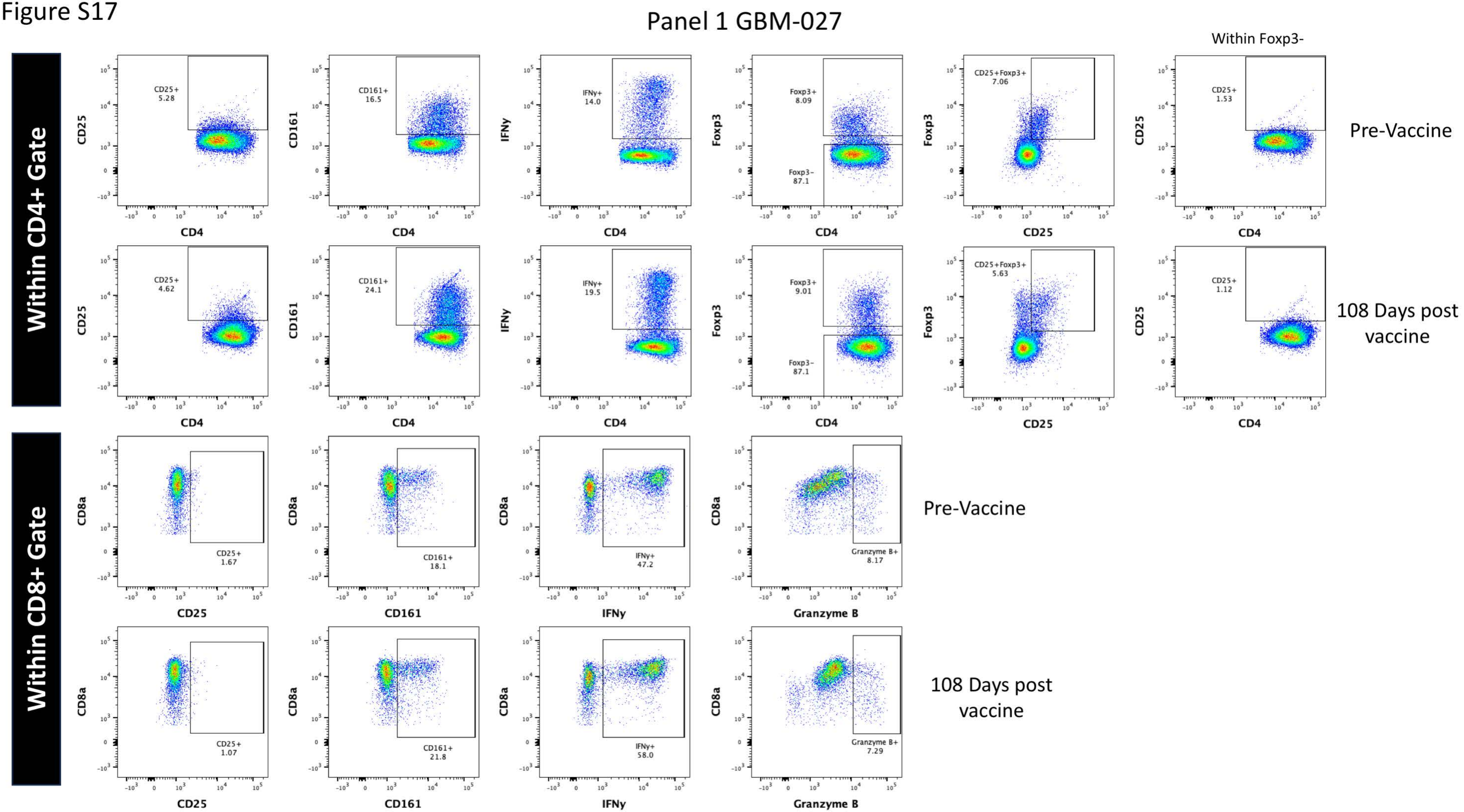
Panel 1 GBM-027
Within CD4+ Gate
Within CD8+ Gate
Within Foxp3-
Pre-Vaccine
108 Days post vaccine
CD25+ 5.28
CD161+ 16.5
IFNy+ 14.0
Foxp3+ 8.09
Foxp3- 87.1
CD25+Foxp3+ 7.06
CD25+ 1.53
CD25+ 4.62
CD161+ 24.1
IFNy+ 19.5
Foxp3+ 9.01
Foxp3- 87.1
CD25+Foxp3+ 5.63
CD25+ 1.12
CD25+ 1.67
CD161+ 18.1
IFNy+ 47.2
Granzyme B+ 8.17
CD25+ 1.07
CD161+ 21.8
IFNy+ 58.0
Granzyme B+ 7.29
CD4
CD25
CD161
IFNy
Foxp3
CD8a
Granzyme B

Figure S18

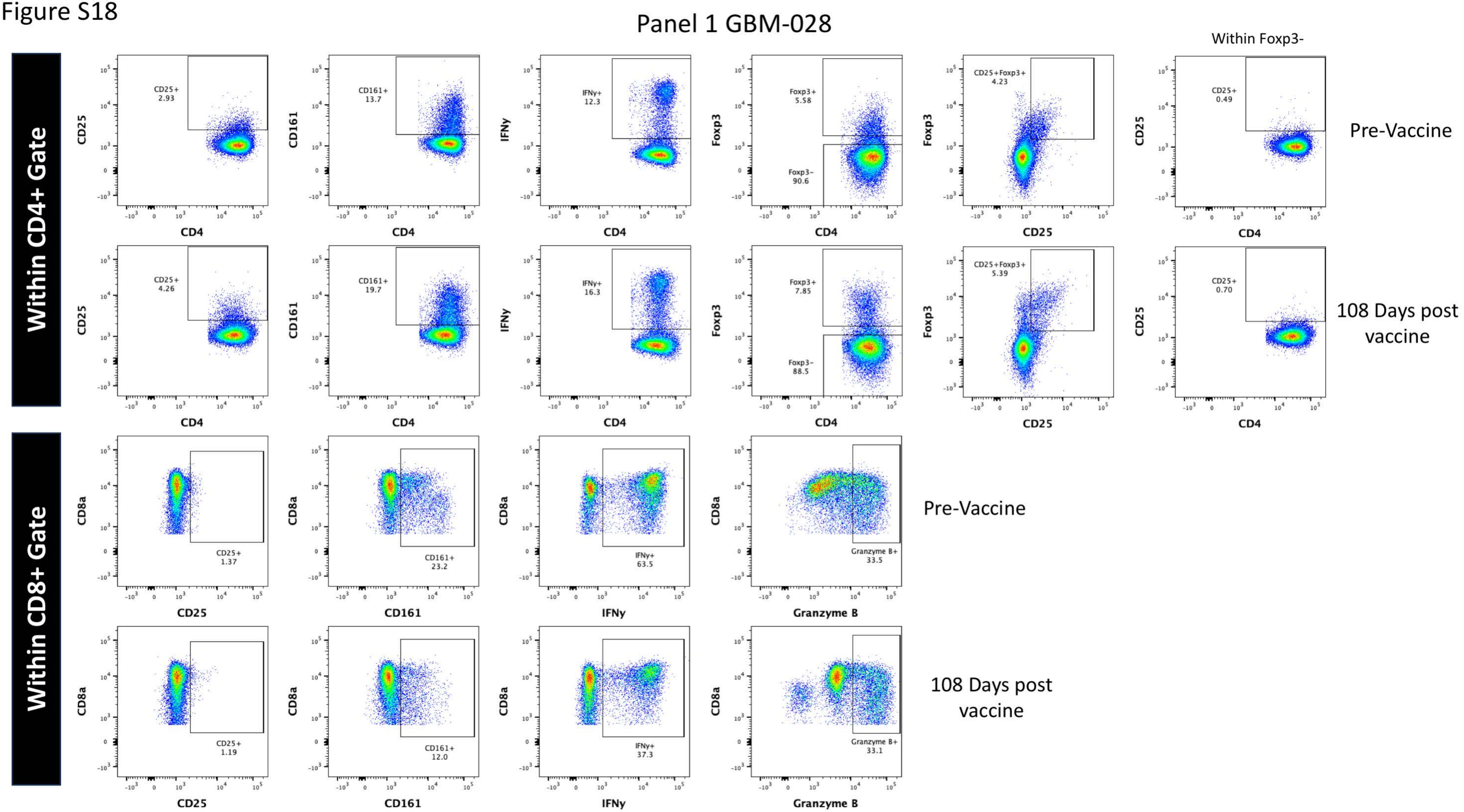
Panel 1 GBM-028
Within CD4+ Gate
Within CD8+ Gate
Within Foxp3-
Pre-Vaccine
108 Days post vaccine
CD25+ 2.93
CD161+ 13.7
IFNy+ 12.3
Foxp3+ 5.58
Foxp3- 90.6
CD25+Foxp3+ 4.23
CD25+ 0.49
CD25+ 4.26
CD161+ 19.7
IFNy+ 16.3
Foxp3+ 7.85
Foxp3- 88.5
CD25+Foxp3+ 5.39
CD25+ 0.70
CD25+ 1.37
CD161+ 23.2
IFNy+ 63.5
Granzyme B+ 33.5
CD25+ 1.19
CD161+ 12.0
IFNy+ 37.3
Granzyme B+ 33.1
CD4
CD25
CD161
IFNy
Foxp3
CD8a
Granzyme B

Figure S19

### Panel 2 Healthy Donor Overall Gating Strategy with Controls

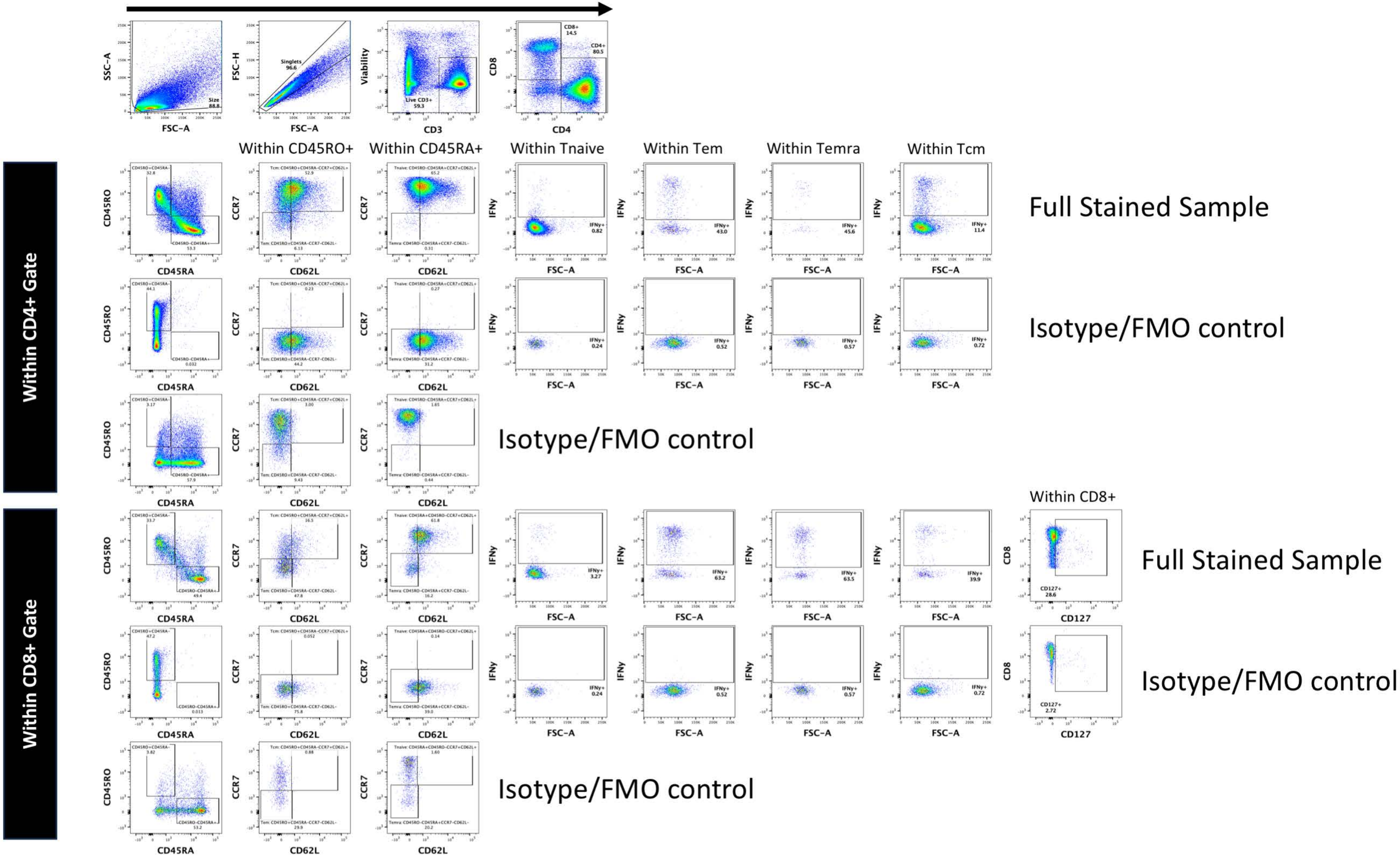

Figure S20

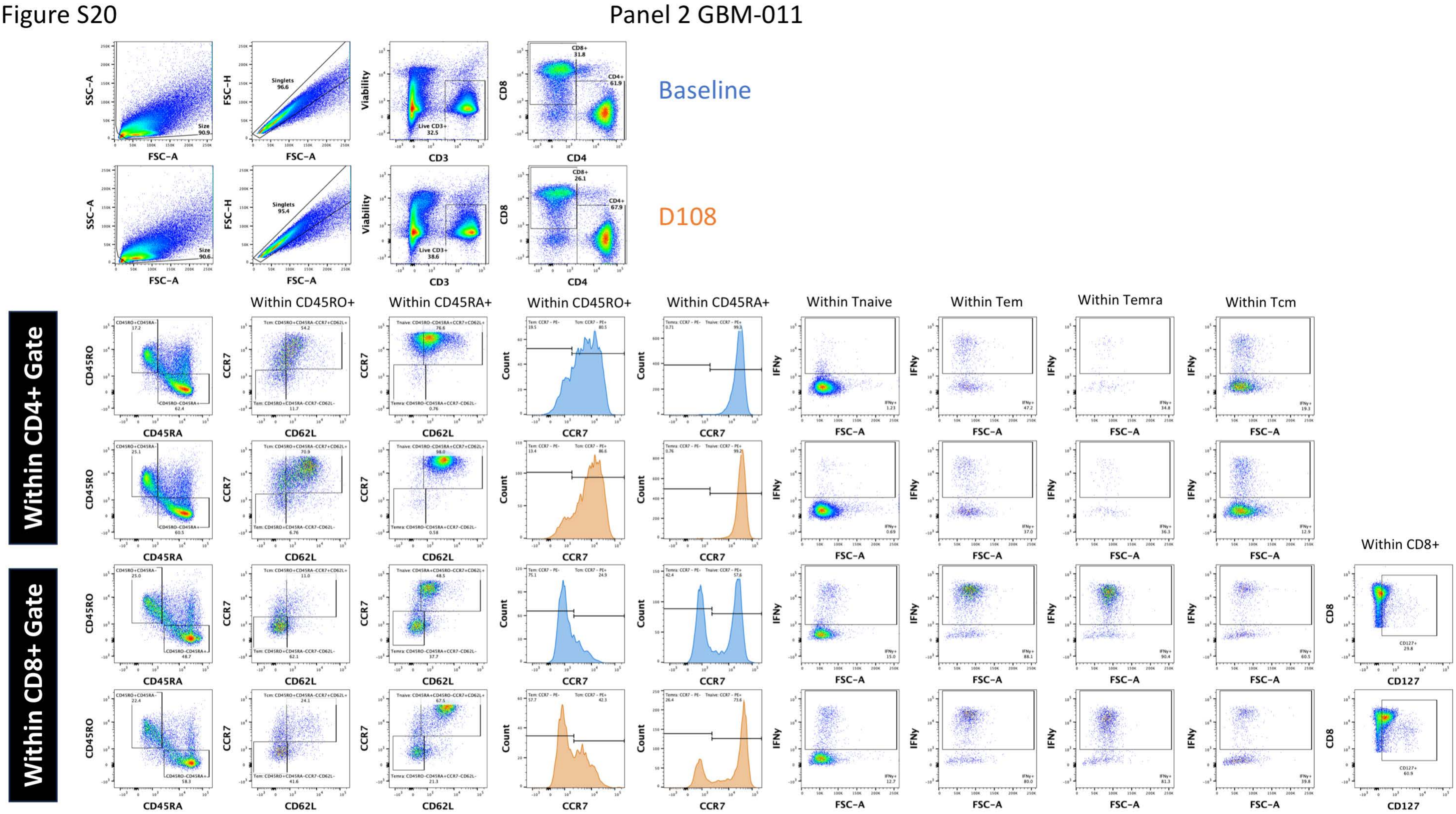

Panel 2 GBM-011
Baseline
D108
Within CD4+ Gate
Within CD8+ Gate
Within CD45RO+
Within CD45RA+
Within CD45RO+
Within CD45RA+
Within Tnaive
Within Tem
Within Temra
Within Tcm
Within CD8+

Figure S21

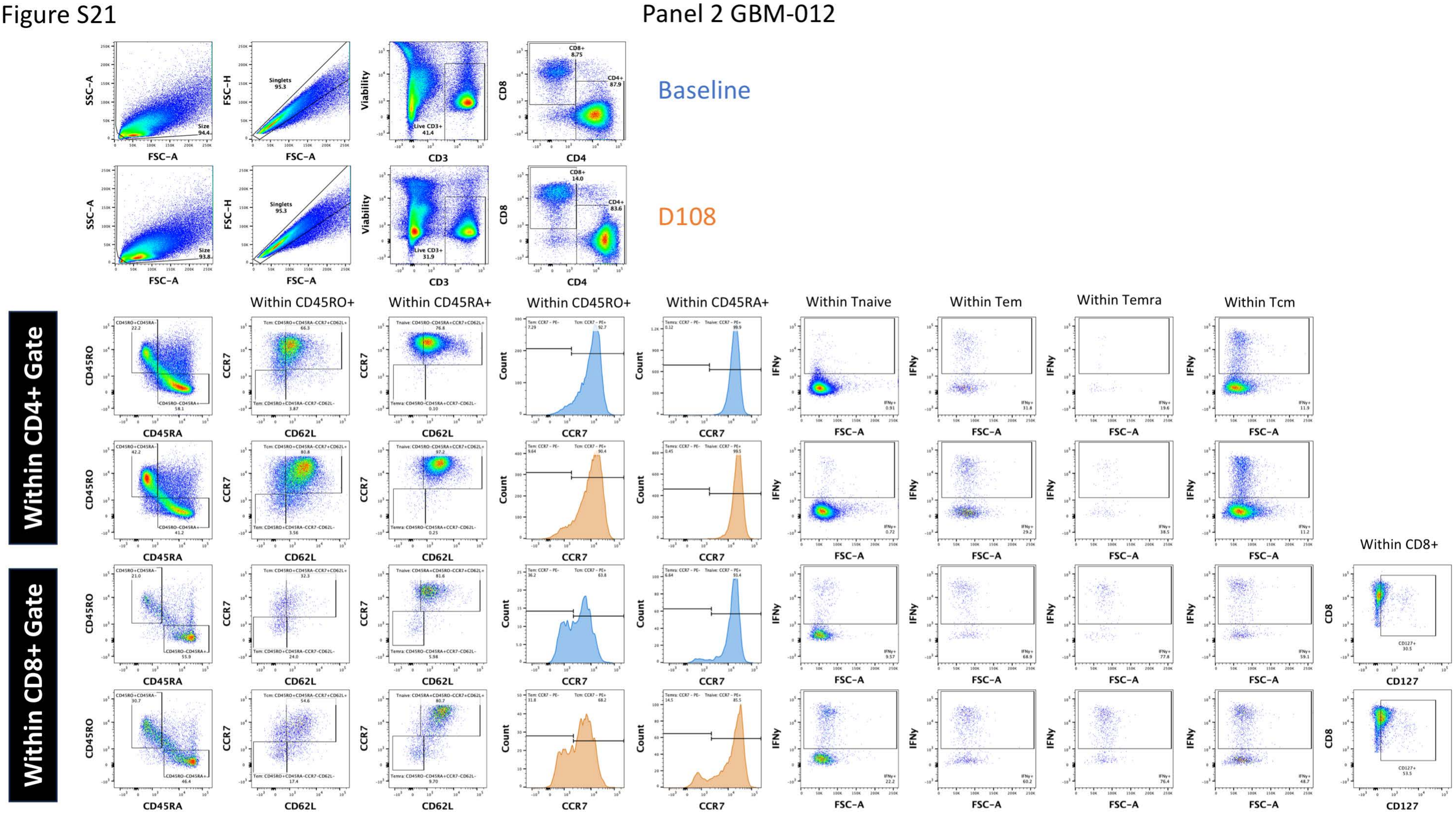
Panel 2 GBM-012
Baseline
D108
Within CD4+ Gate
Within CD8+ Gate
Within CD45RO+
Within CD45RA+
Within CD45RO+
Within CD45RA+
Within Tnaive
Within Tem
Within Temra
Within Tcm
Within CD8+

Figure S22

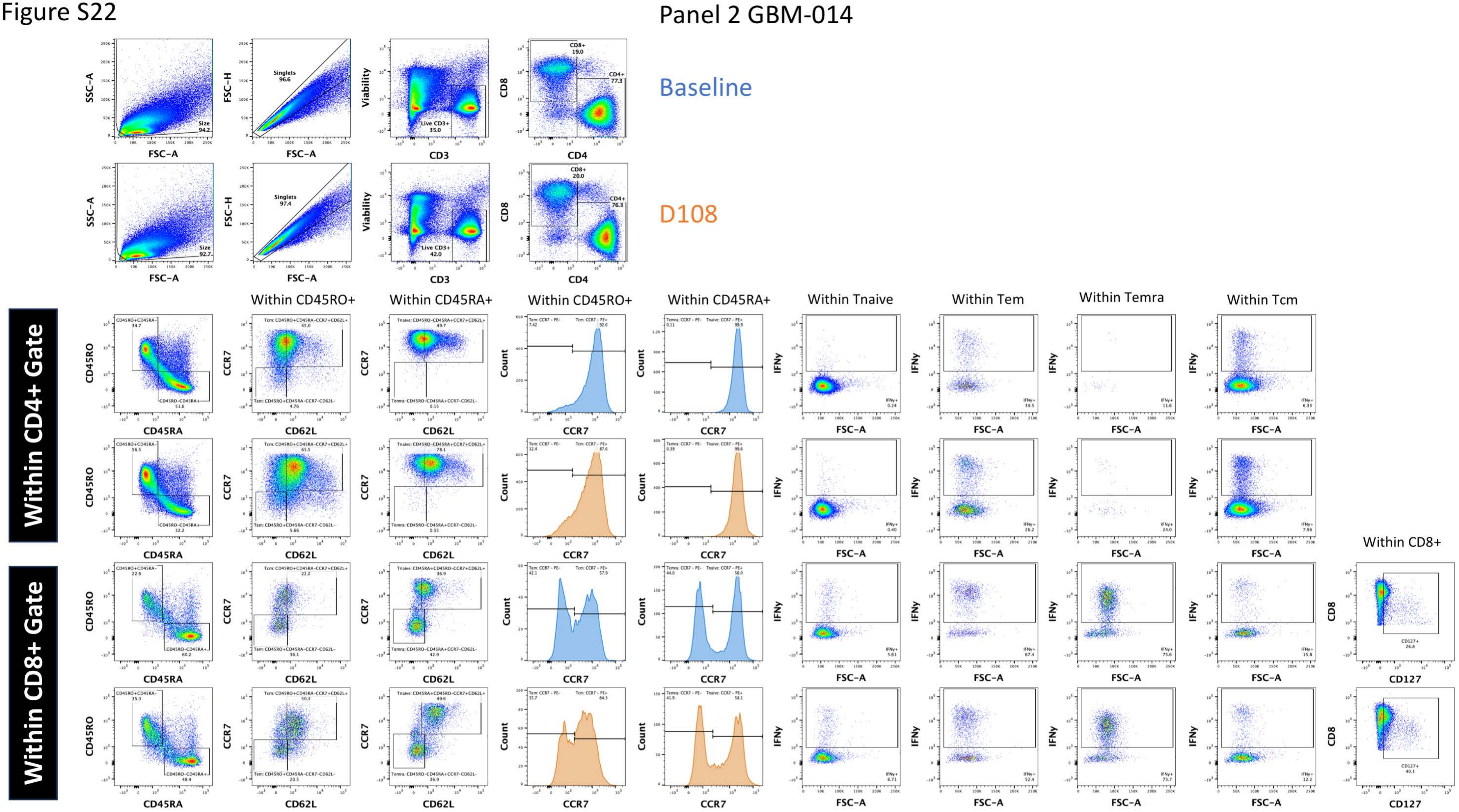
Panel 2 GBM-014
Baseline
D108
Within CD4+ Gate
Within CD8+ Gate
Within CD45RO+
Within CD45RA+
Within CD45RO+
Within CD45RA+
Within Tnaive
Within Tem
Within Temra
Within Tcm
Within CD8+
Singlets
96.6
Size
94.2
Live CD3+
35.0
CD8+
19.0
CD4+
77.3
Singlets
97.4
Size
92.7
Live CD3+
42.0
CD8+
20.0
CD4+
76.3
SSC-A
FSC-A
FSC-H
Viability
CD3
CD8
CD4
CD45RO
CD45RA
CCR7
CD62L
Count
IFNγ
CD127

Figure S23

Panel 2 GBM-015

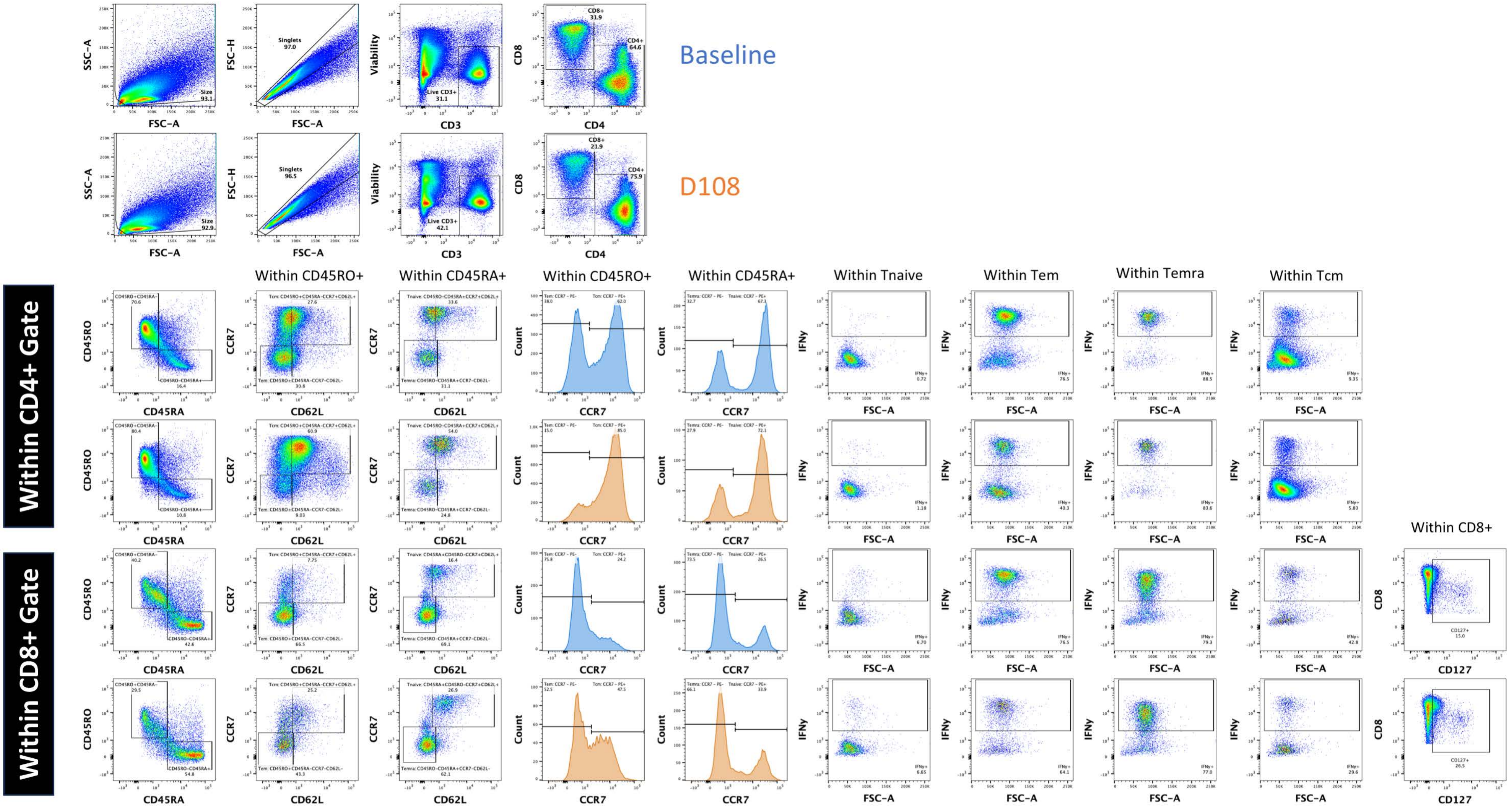

Figure S24

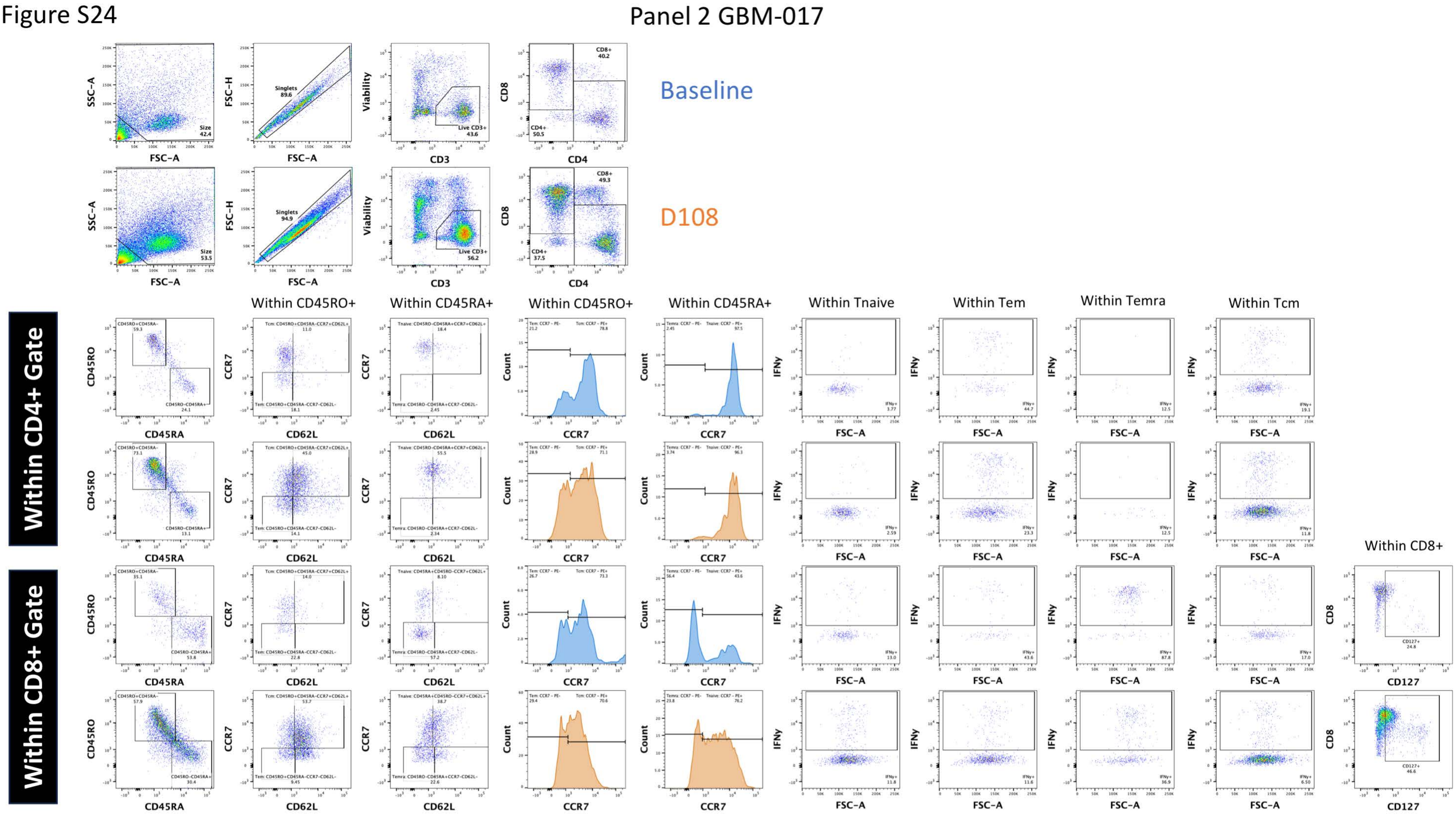
Panel 2 GBM-017
Baseline
D108
Within CD4+ Gate
Within CD8+ Gate
Within CD45RO+
Within CD45RA+
Within CD45RO+
Within CD45RA+
Within Tnaive
Within Tem
Within Temra
Within Tcm
Within CD8+
SSC-A
FSC-A
FSC-H
Viability
CD3
CD8
CD4
CD45RO
CD45RA
CCR7
CD62L
Count
IFNγ
CD127

Figure S25

Panel 2 GBM-018

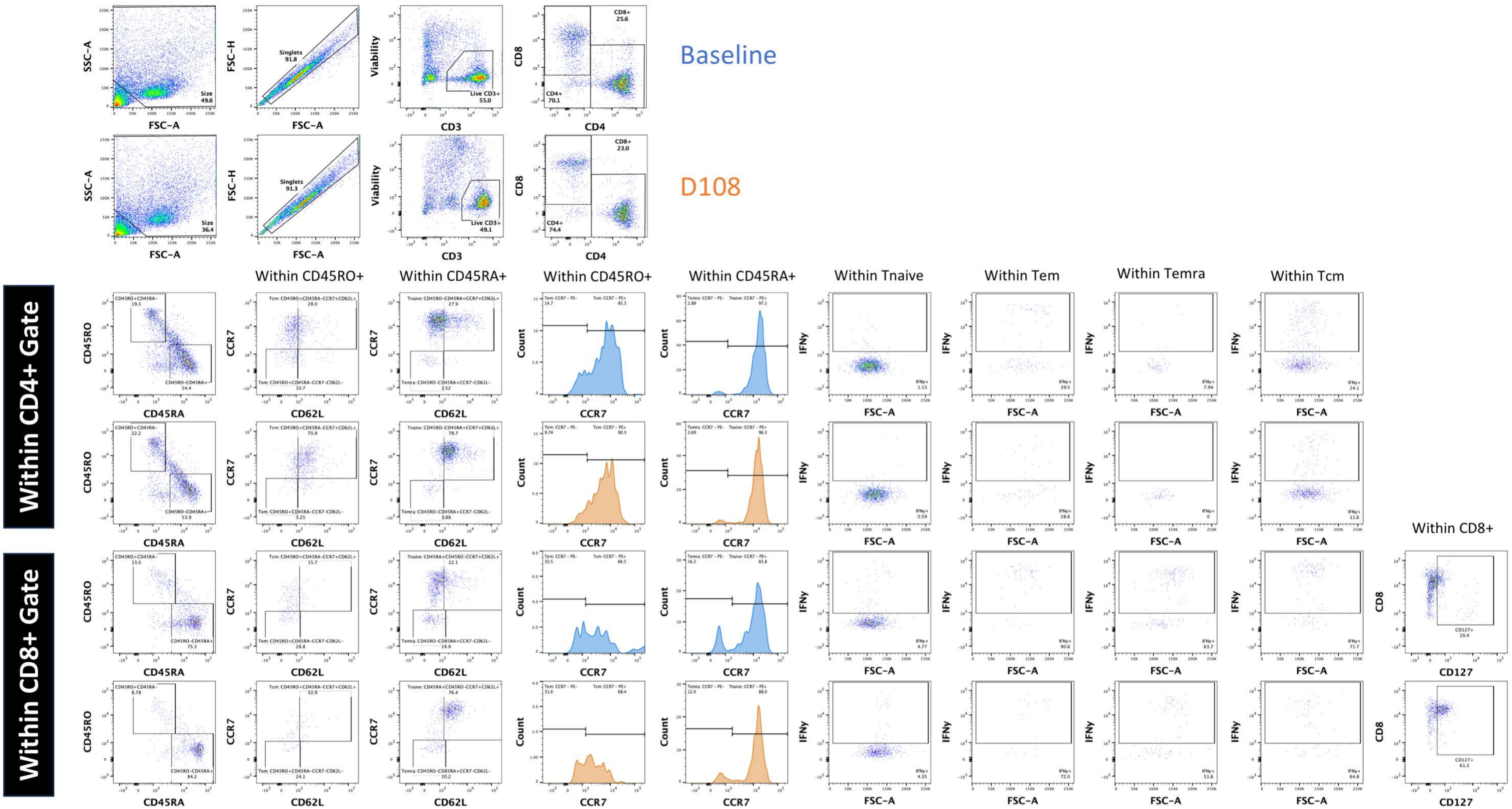

Figure S26

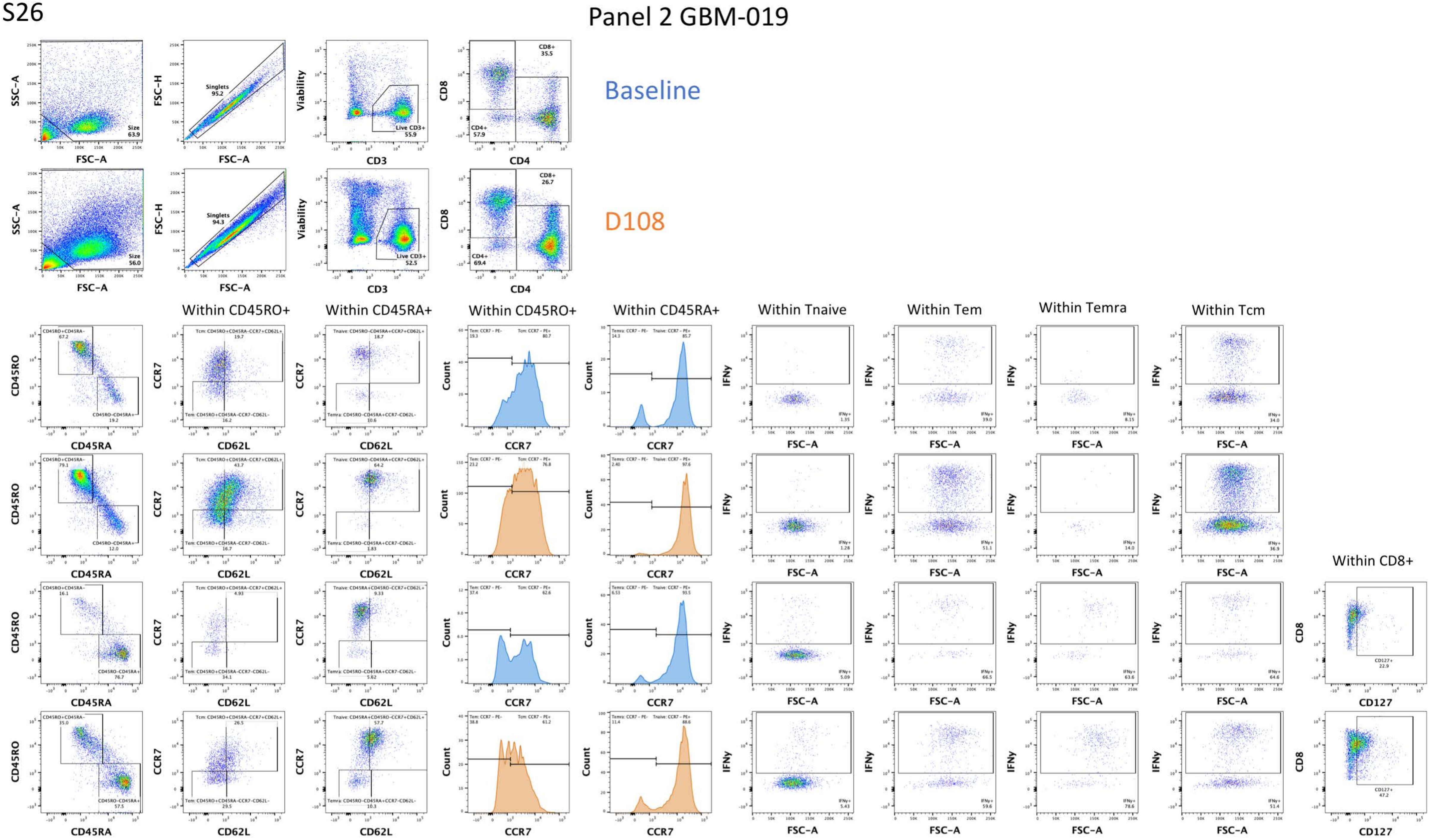

Panel 2 GBM-019
Baseline
D108
Within CD4+ Gate
Within CD8+ Gate
Within CD45RO+
Within CD45RA+
Within CD45RO+
Within CD45RA+
Within Tnaive
Within Tem
Within Temra
Within Tcm
Within CD8+
SSC-A
FSC-A
FSC-H
Viability
CD3
CD8
CD4
CD45RO
CD45RA
CCR7
CD62L
Count
IFNγ
CD127

Figure S27

Panel 2 GBM-021

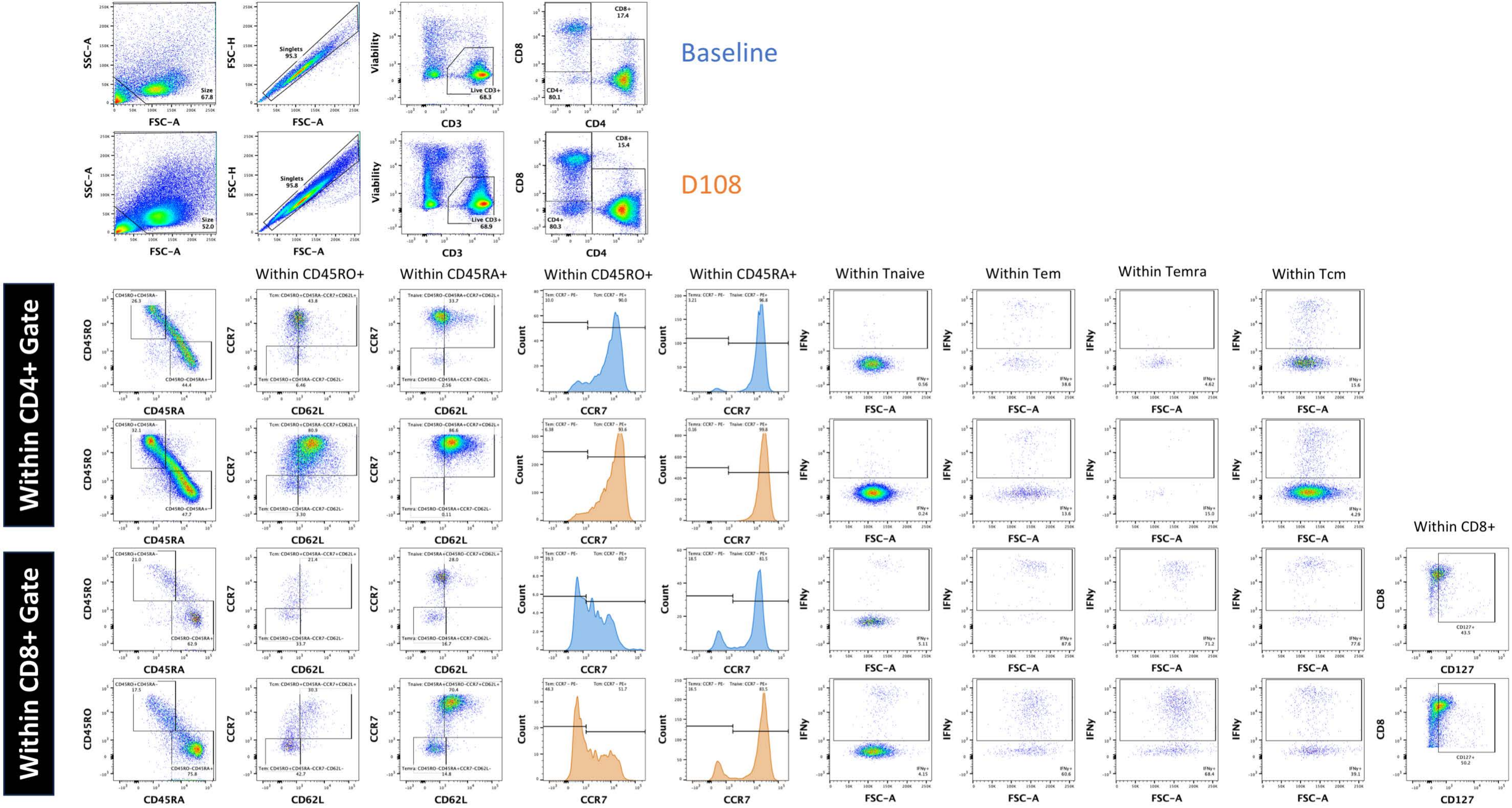

Figure S28

Panel 2 GBM-022

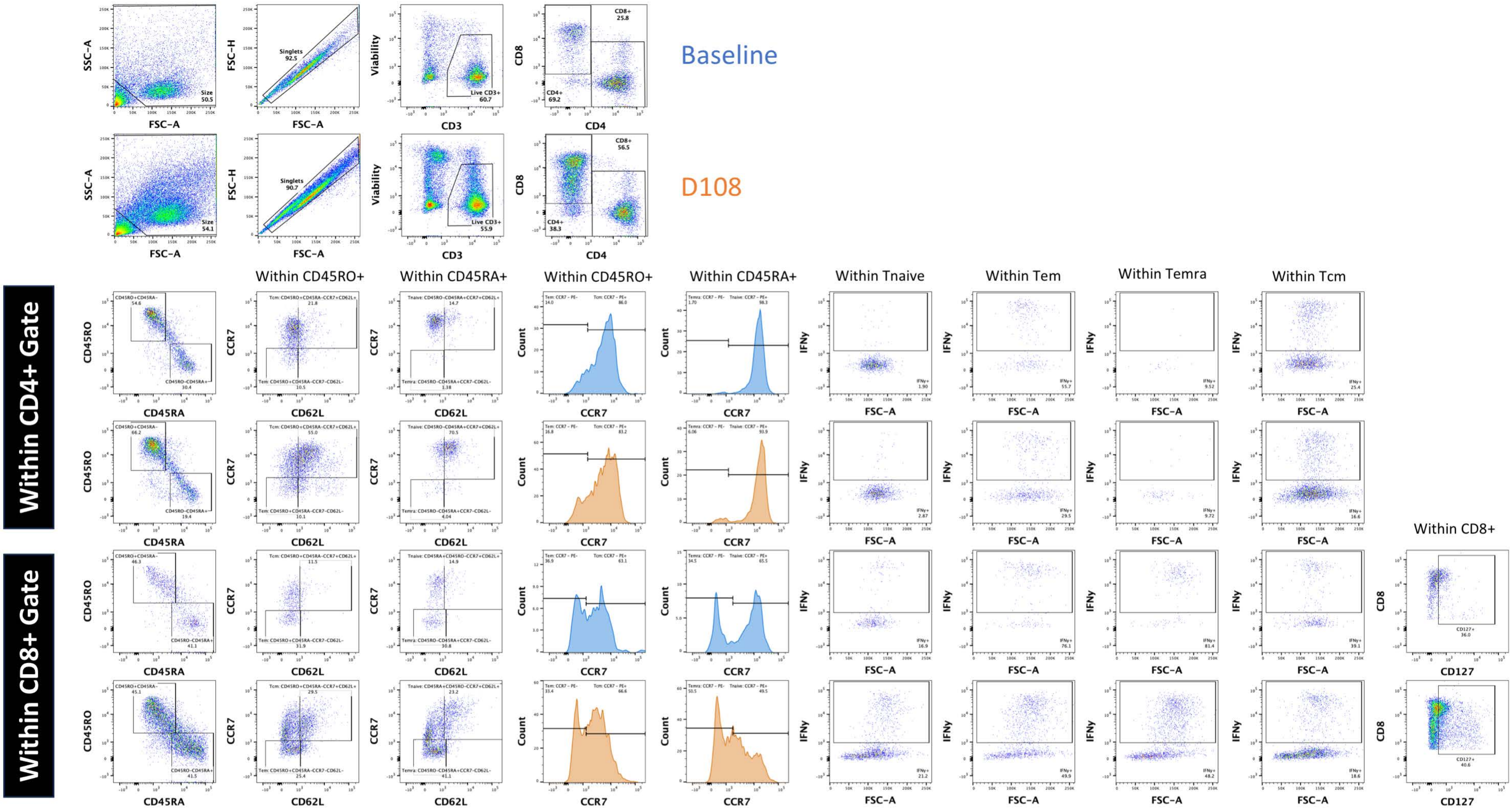

Figure S29

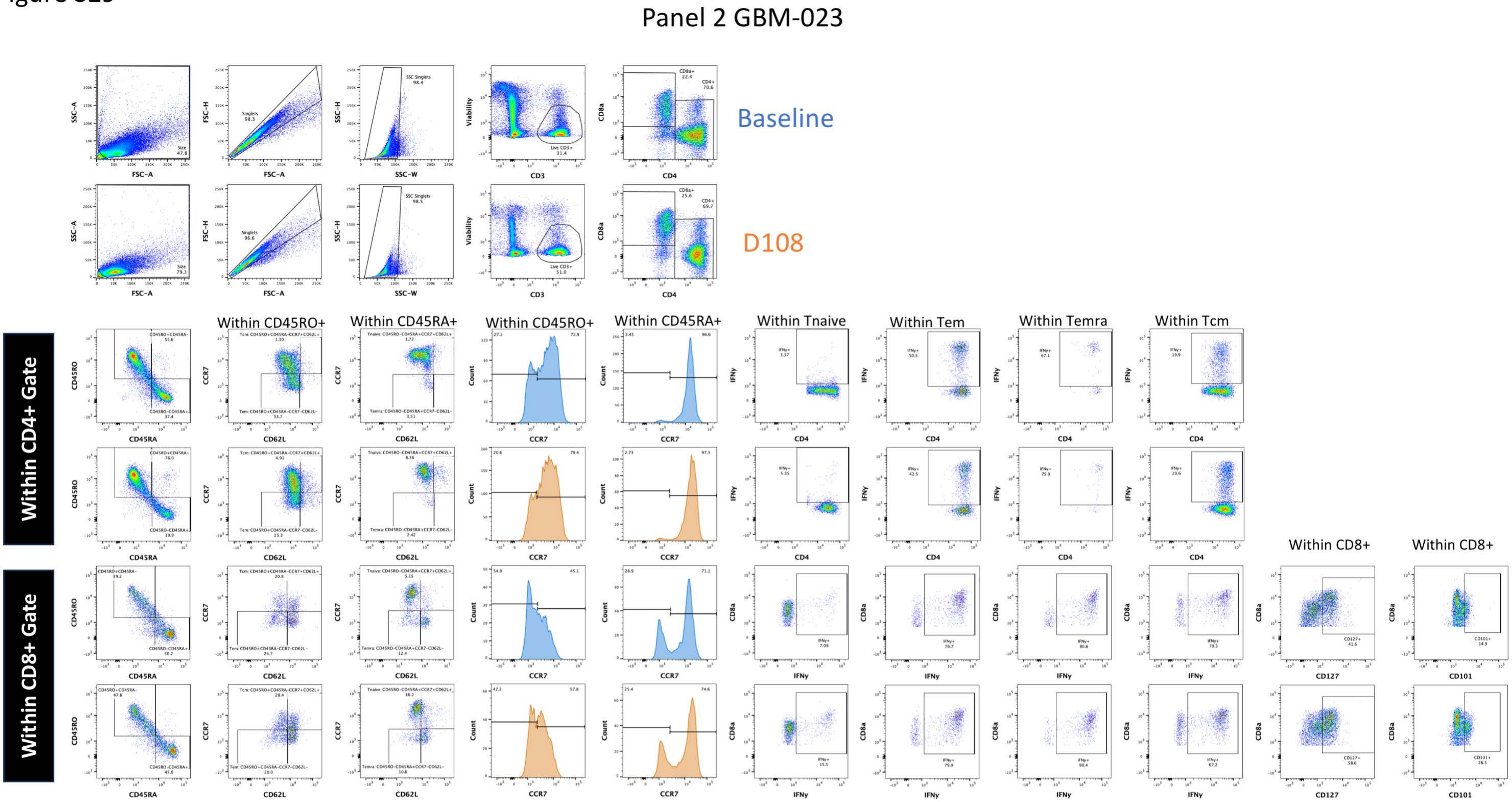
Panel 2 GBM-023
Baseline
D108
Within CD4+ Gate
Within CD8+ Gate
Within CD45RO+
Within CD45RA+
Within CD45RO+
Within CD45RA+
Within Tnaive
Within Tem
Within Temra
Within Tcm
Within CD8+
Within CD8+

Figure S30

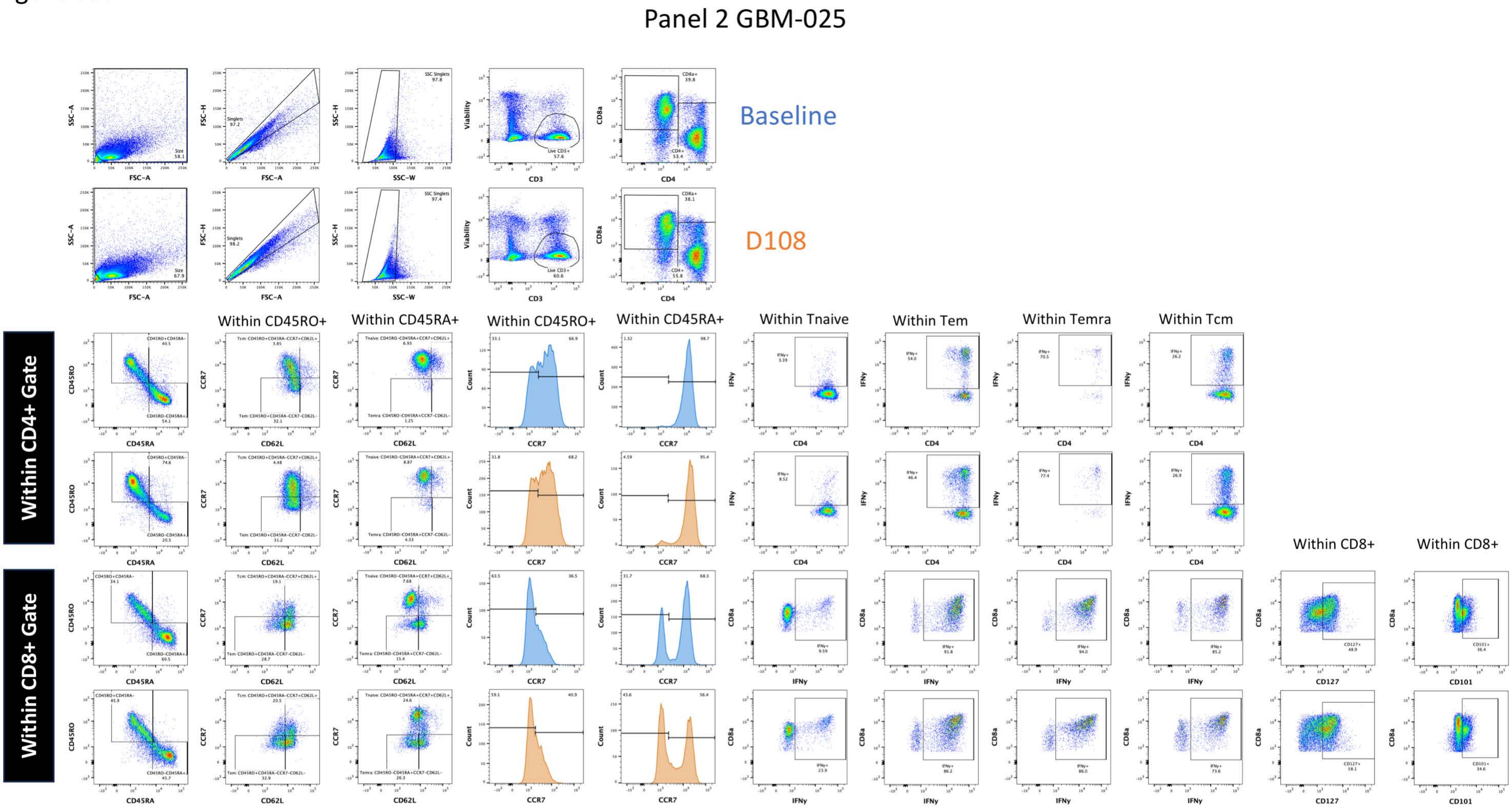

Panel 2 GBM-025
Baseline
D108
Within CD4+ Gate
Within CD8+ Gate
Within CD45RO+
Within CD45RA+
Within CD45RO+
Within CD45RA+
Within Tnaive
Within Tem
Within Temra
Within Tcm
Within CD8+
Within CD8+

Figure S31

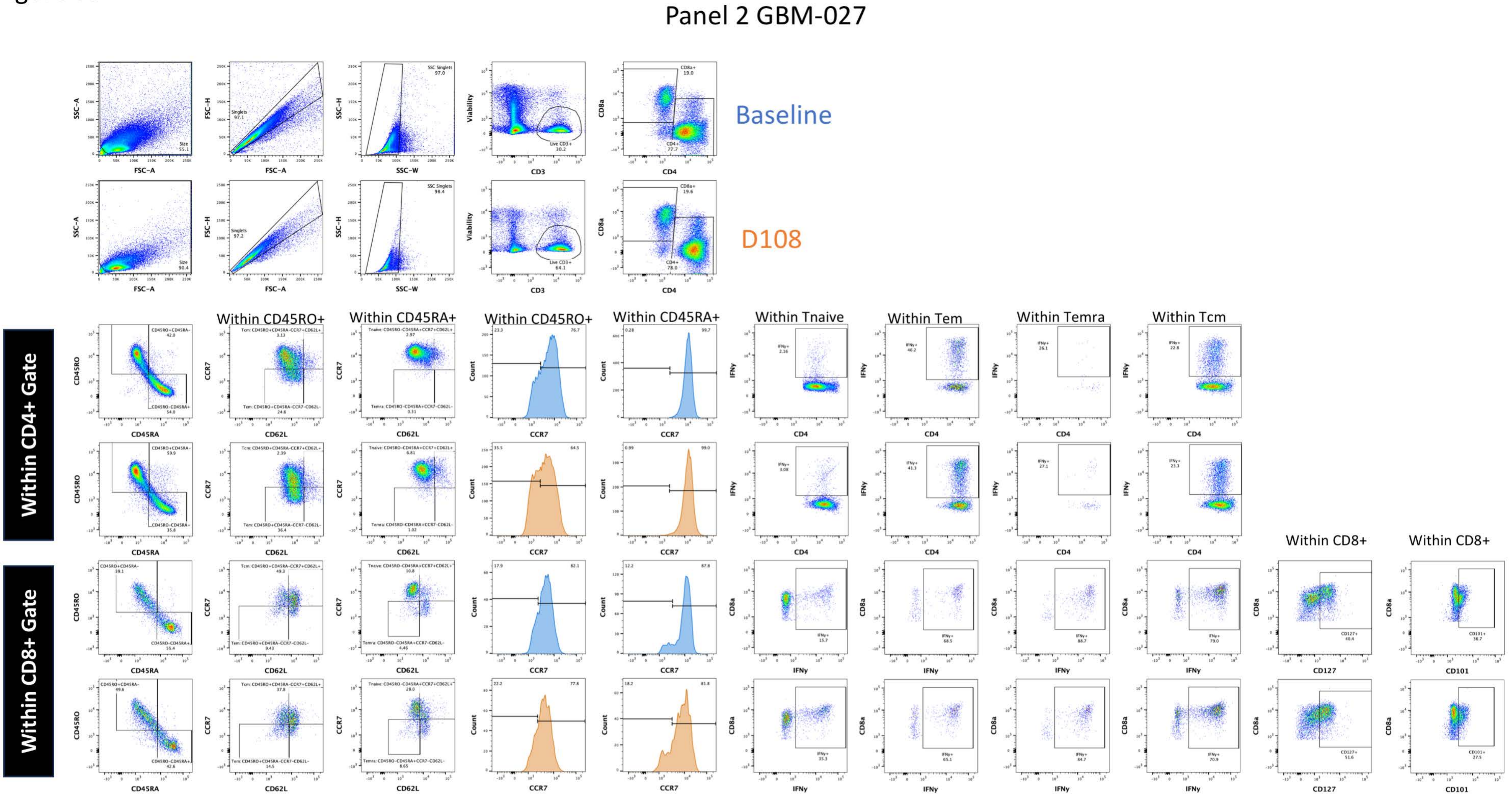
Panel 2 GBM-027
Baseline
D108
Within CD4+ Gate
Within CD8+ Gate
Within CD45RO+
Within CD45RA+
Within CD45RO+
Within CD45RA+
Within Tnaive
Within Tem
Within Temra
Within Tcm
Within CD8+
Within CD8+
FSC-A
SSC-A
FSC-H
SSC-H
SSC-W
CD3
Viability
CD4
CD8a
CD45RA
CD45RO
CD62L
CCR7
Count
IFNγ
CD127
CD101

Figure S32

### Panel 2 GBM-028

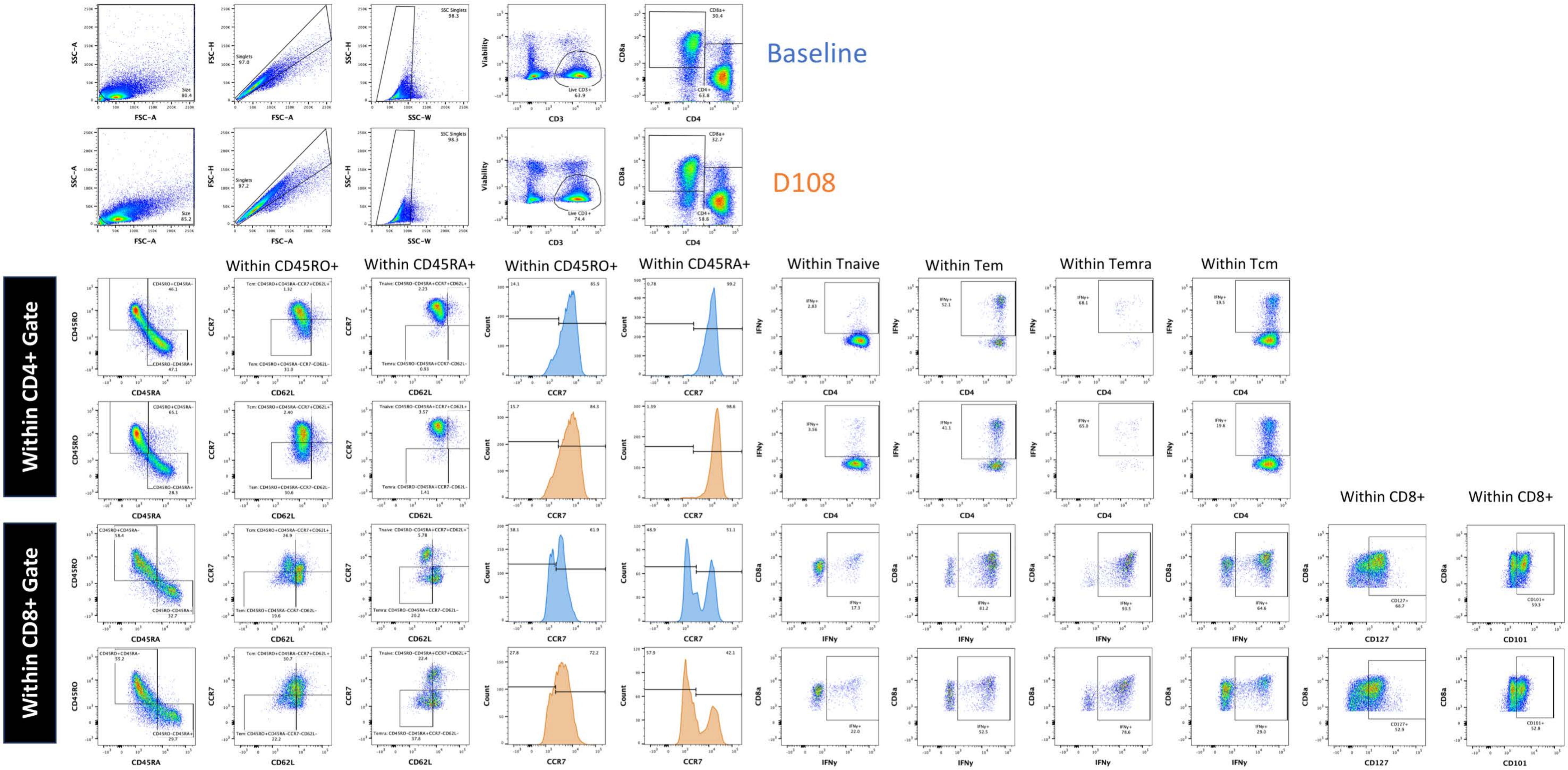
